# Altered anterior default mode network dynamics in progressive multiple sclerosis

**DOI:** 10.1101/2020.11.26.20238923

**Authors:** Giulia Bommarito, Anjali Tarun, Younes Farouj, Maria Giulia Preti, Maria Petracca, Amgad Droby, Mohamed Mounir El Mendili, Matilde Inglese, Dimitri Van De Ville

**Affiliations:** Institute of Bioengineering, Center for Neuroprosthetics, Ecole Polytechnique Fédérale de Lausanne, Geneva, Geneva 1202, Switzerland; Department of Radiology and Medical Informatics, Faculty of Medicine, University of Geneva, Geneva, Geneva 1206, Switzerland; Department of Neuroscience, Rehabilitation, Ophthalmology, Genetics, Maternal and Child Health (DINOGMI), University of Genoa, Genoa, Italy; Department of Neurology, Icahn School of Medicine at Mount Sinai, New York, NY, USA; Ospedale Policlinico San Martino, IRCCS, Largo Daneo 3, 16100, Genoa, Italy

**Keywords:** progressive multiple sclerosis, resting-state functional MRI, innovation-driven co-activation patterns, default mode network, clinical disability

## Abstract

**Background:** Modifications in brain function remain relatively unexplored in progressive multiple sclerosis (PMS), despite their potential to provide new insights into the pathophysiology of this disease stage.

**Objectives:** To characterize the dynamics of functional networks at rest in patients with PMS, and the relation with clinical disability.

**Methods:** Thirty-two patients with PMS underwent clinical and cognitive assessment. The dynamic properties of functional networks, retrieved from transient brain activity, were obtained from patients and 25 healthy controls (HC). Sixteen HC and 19 patients underwent a one-year follow-up clinical and imaging assessment. Differences in the dynamic metrics between groups, their longitudinal changes, and the correlation with clinical disability were explored.

**Results:** PMS patients, compared to HC, showed a reduced dynamic functional activation of the anterior default mode network (aDMN) and its opposite-signed coactivation with the executive-control network, at baseline and follow-up. Processing speed and visuo-spatial memory negatively correlated to aDMN dynamic activity. The anti-coupling between aDMN and auditory/sensory-motor network, temporal-pole/amygdala or salience networks were differently associated to separate cognitive domains.

**Conclusion:** Patients with PMS presented an altered aDMN functional recruitment and anti-correlation with ECN. The aDMN dynamic functional activity and interaction with other networks explained cognitive disability.

## Introduction

Progressive multiple sclerosis (PMS) is characterized by a gradual worsening in clinical disability, occurring from the onset of the disease in the case of primary progressive (PP) form, or after a relapsing-remitting (RR) phase in secondary progressive (SP) multiple sclerosis (MS). PP and SP MS phenotypes are considered different aspects of a unique spectrum^1^ and this disease stage, compared to RR MS, presents with a slower, chronic, compartmentalized inflammation and a prominent neurodegeneration, together with a depleted ability to respond to damage. The contribution of imaging to the definition of PMS ranges from the identification of biomarkers, as atrophy, to the detection, in vivo, of hallmarks such as cortical lesions and diffuse white matter damage^1,2^. Nonetheless, while a growing number of reports is revealing the clinical significance of the networks function disruption occurring in MS, studies targeting brain functional activity in PMS are relatively lacking.

Previous studies of brain function at rest in patients with PMS reported altered network activity or connectivity involving the default mode network (DMN)^3^, attentional, executive control and sensory-motor networks^4–6^. However, the discordant results and the limited number of studies does not allow a unique interpretation of the changes in functional networks occurring in this phase of the disease and the hypothesis of an exhaustion of plasticity characterizing PMS is mainly inferred by studies on RR patients at later stages. Moreover, to our knowledge, only a longitudinal study focusing on the sensory-motor functional network has been performed, so far^7^. As emerging for other neurodegenerative conditions, defining the functional disruption occurring in PMS could contribute to the understanding of pathophysiology, to extricate the changes due to the disease or to brain aging and to pinpoint potential treatments^8^.

Beside conventional analysis, new approaches have recently emerged to explore the time-varying nature of brain functional activity at rest^9^. In healthy subjects, the investigation of the dynamic properties of rs-FC has revealed its ability in better grasping behavioral aspects, compared to the classic time-averaged connectivity^10^. The dynamics of rs-FC have also been investigated in patients at the earlier stages of MS, emphasizing the complex functional changes occurring in this disorder, and their relationship to the cognitive status^11–16^. Moreover, the ability of dynamic rs-FC in better capturing cognitive decline has been showed also in neurodegenerative disorders^17^.

Multiple methodologies have been developed to study the variations over time in functional brain activity. Among them, the characterization of patterns of functional co-activation among regions has been demonstrated able to tackle the pathophysiological processes occurring in neurological or neuropsychiatric disorders, and to parallel the clinical improvement after therapeutic procedures^18–20^

In this study, we hypothesized that the dynamics of functional networks, investigated by means of co-activation patterns, would be altered in patients with PMS and would explain clinical disability. Specifically, we investigated: i) the dynamics of functional networks in patients with PMS, compared to healthy controls (HC); ii) their changes after one-year of follow-up (FU), and iii) their role in explaining cognitive impairment, assessed with the Brief International Cognitive Assessment for Multiple Sclerosis, and clinical disability.

## Materials and Methods

### Participants

Forty-eight patients with either PP or SP MS and 26 healthy controls (HC) were prospectively recruited. Inclusion and exclusion criteria are detailed in the Supplementary Information (SI). After image quality check and motion correction (detailed in SI), the final sample included 57 subjects: 32 patients with PMS (mean age: 50.8±9.9 years, 20 women, 18 patients with PP MS and 14 patients with SP MS) and 25 HC (mean age: 47.3±8.7 years, 10 women). All subjects underwent MRI and, on the same day, patients underwent clinical evaluation, including expanded disability status scale (EDSS), timed-25-foot-walking (T25FW) test, 9-hole-peg-test (9HPT), and neuropsychological assessment with the symbol digit modalities test (SDMT), California verbal learning test second edition (CVLT-II) and the brief visuospatial memory test revised (BVMT-R). For the CVLT-II and the BVMT-R the first five or three recall trials were considered, respectively. Raw scores were transformed into Z-scores according to^21^. Impairment for a single test was assessed on the level of 1.5 SD, and patients were defined as cognitively impaired (CI) or preserved (CP) based on scoring outside the normal range in one or more of the tests^22^.

Thirty-five (16 HC and 19 patients, 11 with PP MS and 8 with SP MS) out of the 57 subjects included at baseline underwent MRI and clinical evaluation after 1-year ([mean ±SD]= 11±2 months). Subjects who were included at FU or dropped out from the study did not differ in terms of demographic, clinical and imaging variable (see Table SI 1).

The study was approved by the Institutional Review Board of the Icahn School of Medicine at Mount Sinai, and all the subjects gave written informed consent to participate.

### MRI acquisition

All the participants underwent MRI on the same 3.0□T scanner (Magnetom Skyra, Erlangen, Siemens, Germany) using a 32-channel head coil, with a constant protocol at baseline and FU, including T2-weighted, T1-weighted, and a single shot gradient echo planar imaging sequence for the resting state. Sequence parameters are reported in the SI.

### Structural MRI analysis

T2 and T1 lesions were segmented, and lesion volumes (T2 LV and T1 LV, respectively) obtained using a semiautomatic technique (Jim 7, Xinapse Systems, Northants, UK). The T1-weighted images of PMS patients underwent a lesion in-painting procedure prior to the brain extraction and anatomical structures’ segmentation^23^. The baseline normalized brain volume (BV), grey matter (GMV) and white matter volumes (WMV), and the FU percentage brain volume changes were estimated using SIENAX and SIENA, respectively^24^.

### Functional resting state MRI preprocessing and iCAPs analysis

Resting state functional magnetic resonance imaging (rs-fMRI) data were preprocessed using SPM12 (FIL, UCL, UK), complementary functions of the Data Processing Assistant for Resting-State fMRI (DPARSF)^25^ and the Individual Brain Atlases using Statistical Parametric Mapping toolboxes^26^. Preprocessing steps included: realignment and discarding of the first 10 volumes, resulting in 390 time-points, co-registration of the T1-weighted and functional images, segmentation of the structural images into white matter (WM), grey matter and cerebrospinal fluid (CSF). The regression of the nuisance variables, including the average WM and CSF signals and six motion parameters, was performed using DPARSF toolbox. A spatial smoothing (Gaussian kernel of a 6 mm full width at half maximum) was finally applied.

The innovation-driven-coactivation-patterns (iCAPs) analysis is a novel technique that allows to derive a set of whole-brain spatial patterns of regions whose activity simultaneously increases or decreases, thus characterized by similar functional dynamic behavior. These are obtained by detecting the *transients* or changes in rs-fMRI activity time-courses and each iCAP represents a brain functional *state* or network. The procedure was performed using a publicly available implementation (https://c4science.ch/source/iCAPs/). For a comprehensive explanation of the methodology, please refer to^27^, to SI, and Figure 1. The iCAPs retrieved from the baseline sample were fitted into the FU sample, as detailed in the SI. From now on, the terms “iCAPs”, “networks” or “states” will be used interchangeably to describe the spatial patterns of functional activations derived from the iCAPs analysis.

**Figure 1.**
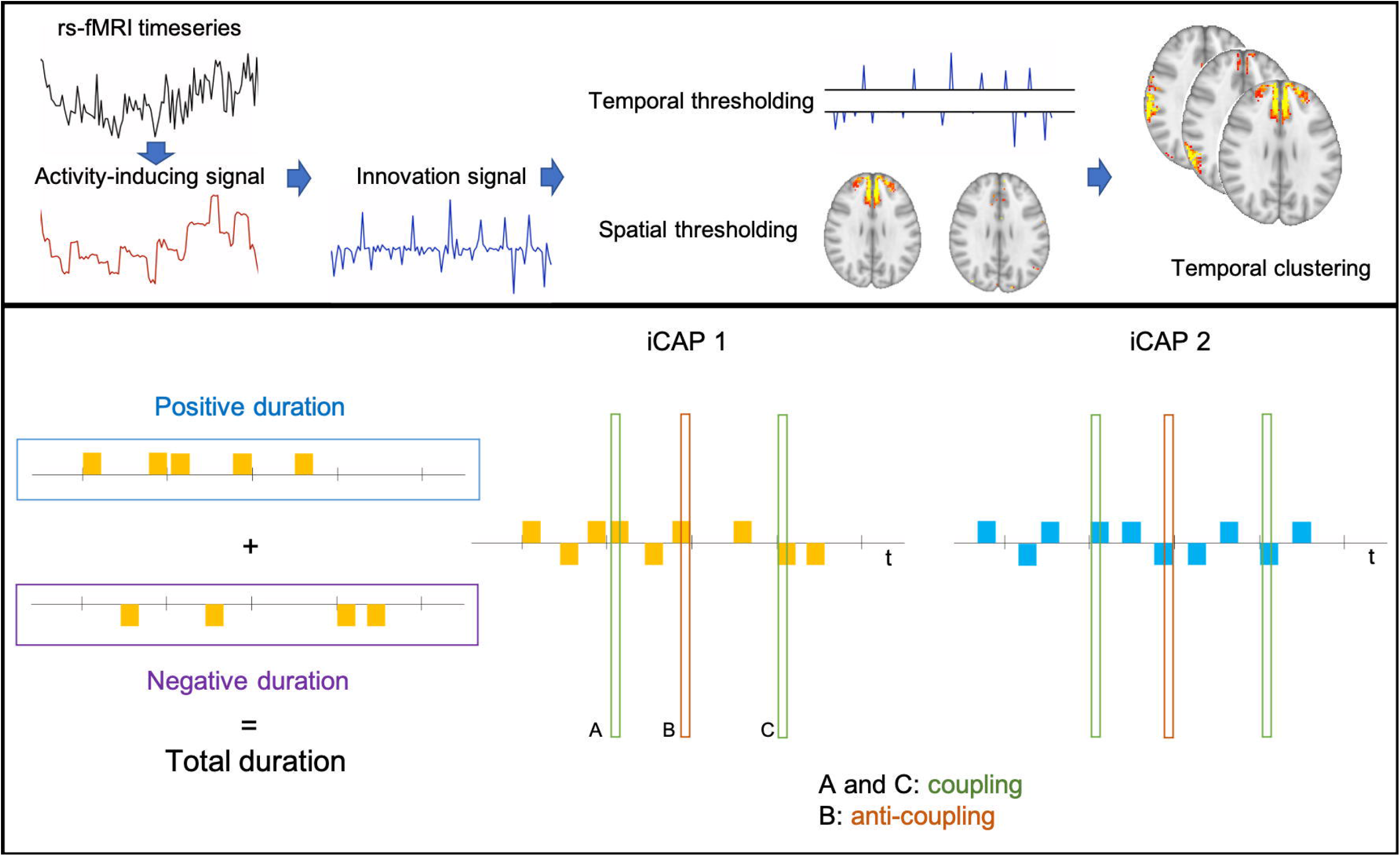
Top: fMRI time-courses undergo a deconvolution step to obtain activity-inducing signals. Innovation signals, obtained by computing the temporal derivative of the activity-inducing signals, enter a two-step thresholding procedure (temporal and spatial) and the resulting frames undergo the temporal clustering step to extract iCAPs. Bottom: the total duration for each iCAP was computed considering both the positive and negative durations, i.e. the time-points when that iCAP was active (after thresholding using a cut-off of |1|). The overlap between 2 iCAPs was measured with the jaccard score, obtained dividing the number of time-points when the two iCAPs were simultaneously active for the number of time-points when at least one of the two iCAPs was active. The couplings and anti-couplings represented the time-points when the 2 iCAPs co- (de)activated (A and C) or showed an opposite co(de)-activation (B), respectively.

### iCAPs temporal properties

For each subject at baseline and FU, we obtained iCAPs’ temporal characteristics: i) the *duration* of overall functional activation, a metric describing the amount of sustained activity (or functional “engagement”) for that specific iCAP; for each pair of iCAPs ii) the *coupling*: same-signed coactivation, and iii) the *anti-coupling*: opposite-signed coactivation (Figure 1). Couplings and anti-couplings reflect the amount of overlap between the activity, in the same or the opposite signs respectively, of each iCAPs pair, and are closely related to the networks’ static functional connectivity.

### Statistical analysis

Demographic and clinical parameters were compared using unpaired and paired t-test between HC and patients with PMS or between values at baseline and FU, respectively.

Analysis of covariance (ANCOVA) was used to compare iCAPs duration between HC and PMS patients and between CI and CP patients, using age, gender and education as covariates. A false discovery rate (FDR) correction according to^28^ was applied. To compare FU and baseline iCAPs temporal properties, a paired t-test within each group and a repeated measure ANCOVA analysis, considering the group x time interaction, were used. Further analyses on coupling/anti-coupling features were computed only for iCAPs whose duration was significantly different in PMS patients compared to HC. ICAPs temporal metrics were correlated with T2 and T1 LV using Spearman’s rank correlation coefficient.

Partial least squares correlation (PLSC) analysis was used to investigate patterns of correlation between clinical parameters and the temporal properties of iCAPs. Age, gender and education were stepwise regressed from each variable, except for the normalized cognitive tests Z scores. The toolbox used to run PLSC analysis is publicly available (https://miplab.epfl.ch/index.php/software/PLS). Since aDMN temporal properties resulted altered in patients with PMS compared to HC (see Results), we assessed their relationship with clinical disability. Grouped PLSC analyses were performed to assess the different contribution of aDMN metrics to cognitive test scores in CI or CP patients. Moreover, we performed an exploratory analysis to assess any relationship between all iCAPs durations and clinical test scores.

## Data availability

Data supporting the findings of this study will be shared upon request.

## Results

### Demographic and clinical data

Demographic data for the two groups and clinical data for the patients with PMS are reported in Table 1. Table SI 1 reports data for the subjects who underwent FU and drop out subjects. No significant differences in demographic variables were observed between HC and PMS, while BV, WMV and GMV were significantly lower in PMS patients. PMS patients did not differ in terms of any clinical parameter of disability between baseline and FU (Table SI 2). Twenty-four patients were classified as CI at baseline, and the PMS group showed low normalized scores at the BVMT and SDMT tests (see also SI and Figure SI 3).

**Table 1.**
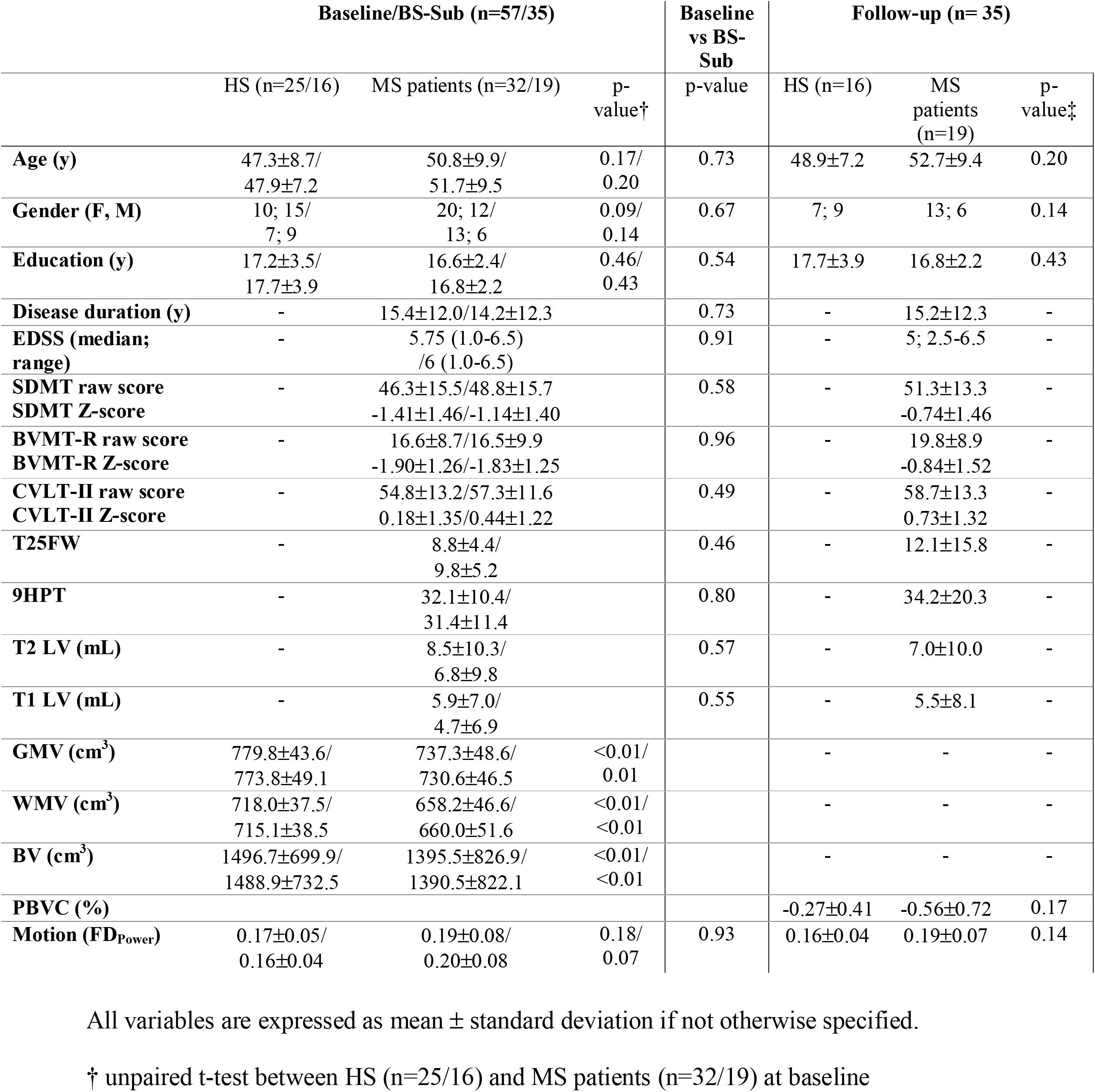
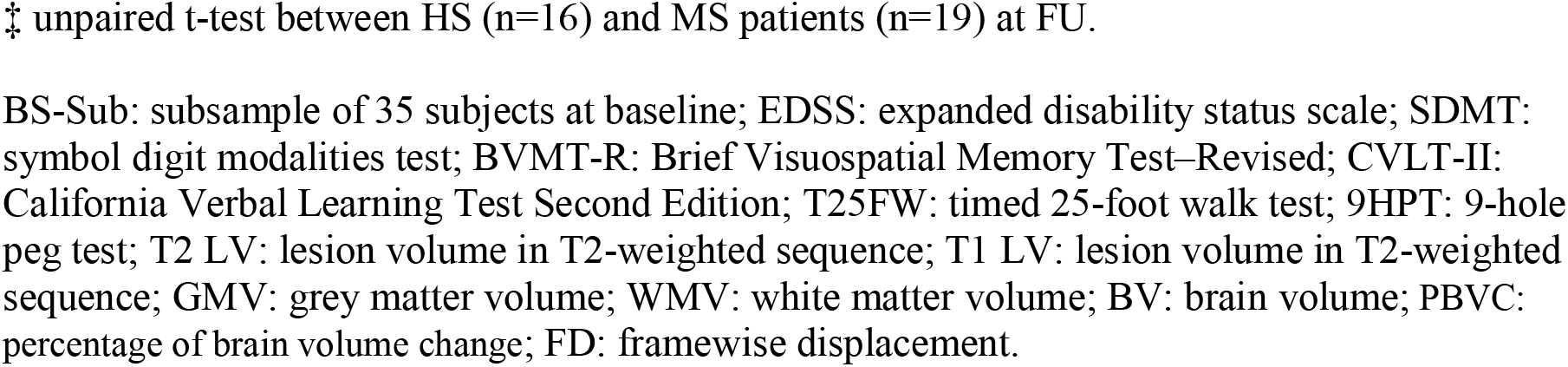
Demographic and clinical data at baseline and follow-up.

|  | Baseline/BS-Sub (n=57/35) |  |  | Baseline<br>vs BS-Sub<br>p-value | Follow-up (n= 35) |  |  |
| --- | --- | --- | --- | --- | --- | --- | --- |
|  | HS (n=25/16) | MS patients (n=32/19) | p-value† |  | HS (n=16) | MS<br>patients<br>(n=19) | p-value‡ |
| **Age (y)** | 47.3±8.7/<br>47.9±7.2 | 50.8±9.9/<br>51.7±9.5 | 0.17/<br>0.20 | 0.73 | 48.9±7.2 | 52.7±9.4 | 0.20 |
| **Gender (F, M)** | 10; 15/<br>7; 9 | 20; 12/<br>13; 6 | 0.09/<br>0.14 | 0.67 | 7; 9 | 13; 6 | 0.14 |
| **Education (y)** | 17.2±3.5/<br>17.7±3.9 | 16.6±2.4/<br>16.8±2.2 | 0.46/<br>0.43 | 0.54 | 17.7±3.9 | 16.8±2.2 | 0.43 |
| **Disease duration (y)** | - | 15.4±12.0/14.2±12.3 |  | 0.73 | - | 15.2±12.3 | - |
| **EDSS (median;**<br>**range)** | - | 5.75 (1.0-6.5)<br>/6 (1.0-6.5) |  | 0.91 | - | 5; 2.5-6.5 | - |
| **SDMT raw score** | - | 46.3±15.5/48.8±15.7 |  | 0.58 | - | 51.3±13.3 | - |
| **SDMT Z-score** | - | -1.41±1.46/-1.14±1.40 |  |  | - | -0.74±1.46 |  |
| **BVMT-R raw score** | - | 16.6±8.7/16.5±9.9 |  | 0.96 | - | 19.8±8.9 | - |
| **BVMT-R Z-score** | - | -1.90±1.26/-1.83±1.25 |  |  | - | -0.84±1.52 |  |
| **CVLT-II raw score** | - | 54.8±13.2/57.3±11.6 |  | 0.49 | - | 58.7±13.3 | - |
| **CVLT-II Z-score** | - | 0.18±1.35/0.44±1.22 |  |  | - | 0.73±1.32 |  |
| **T25FW** | - | 8.8±4.4/<br>9.8±5.2 |  | 0.46 | - | 12.1±15.8 | - |
| **9HPT** | - | 32.1±10.4/<br>31.4±11.4 |  | 0.80 | - | 34.2±20.3 | - |
| **T2 LV (mL)** | - | 8.5±10.3/<br>6.8±9.8 |  | 0.57 | - | 7.0±10.0 | - |
| **T1 LV (mL)** | - | 5.9±7.0/<br>4.7±6.9 |  | 0.55 | - | 5.5±8.1 | - |
| **GMV (cm<sup>3</sup>)** | 779.8±43.6/<br>773.8±49.1 | 737.3±48.6/<br>730.6±46.5 | <0.01/<br>0.01 |  | - | - | - |
| **WMV (cm<sup>3</sup>)** | 718.0±37.5/<br>715.1±38.5 | 658.2±46.6/<br>660.0±51.6 | <0.01/<br><0.01 |  | - | - | - |
| **BV (cm<sup>3</sup>)** | 1496.7±699.9/<br>1488.9±732.5 | 1395.5±826.9/<br>1390.5±822.1 | <0.01/<br><0.01 |  | - | - | - |
| **PBVC (%)** |  |  |  |  | -0.27±0.41 | -0.56±0.72 | 0.17 |
| **Motion (FD<sub>Power</sub>)** | 0.17±0.05/<br>0.16±0.04 | 0.19±0.08/<br>0.20±0.08 | 0.18/<br>0.07 | 0.93 | 0.16±0.04 | 0.19±0.07 | 0.14 |

All variables are expressed as mean ± standard deviation if not otherwise specified.
† unpaired t-test between HS (n=25/16) and MS patients (n=32/19) at baseline
‡ unpaired t-test between HS (n=16) and MS patients (n=19) at FU.
BS-Sub: subsample of 35 subjects at baseline; EDSS: expanded disability status scale; SDMT: symbol digit modalities test; BVMT-R: Brief Visuospatial Memory Test–Revised; CVLT-II: California Verbal Learning Test Second Edition; T25FW: timed 25-foot walk test; 9HPT: 9-hole peg test; T2 LV: lesion volume in T2-weighted sequence; T1 LV: lesion volume in T1-weighted sequence; GMV: grey matter volume; WMV: white matter volume; BV: brain volume; PBVC: percentage of brain volume change; FD: framewise displacement.

### Spatial properties of iCAPs

Eleven iCAPs with different spatial distribution were obtained and considered for subsequent analysis. The networks represented in each iCAP are shown in Figure 2 and named based on the observed spatial pattern and its overlap with well-known functional networks (see Table SI 3). Specifically, they represented the deep grey matter (DGM), auditory/sensory-motor (Aud/SM), primary visual (PrimVis), executive control (ECN), anterior DMN (aDMN), salience network (SAL), temporo-parietal/language (TempPar/Lan), secondary visual (SecVis), precuneus/posterior DMN (pCun/pDMN), amygdala/temporal pole (Amy/TP) and orbito-frontal cortex (OFC).

**Figure 2.**
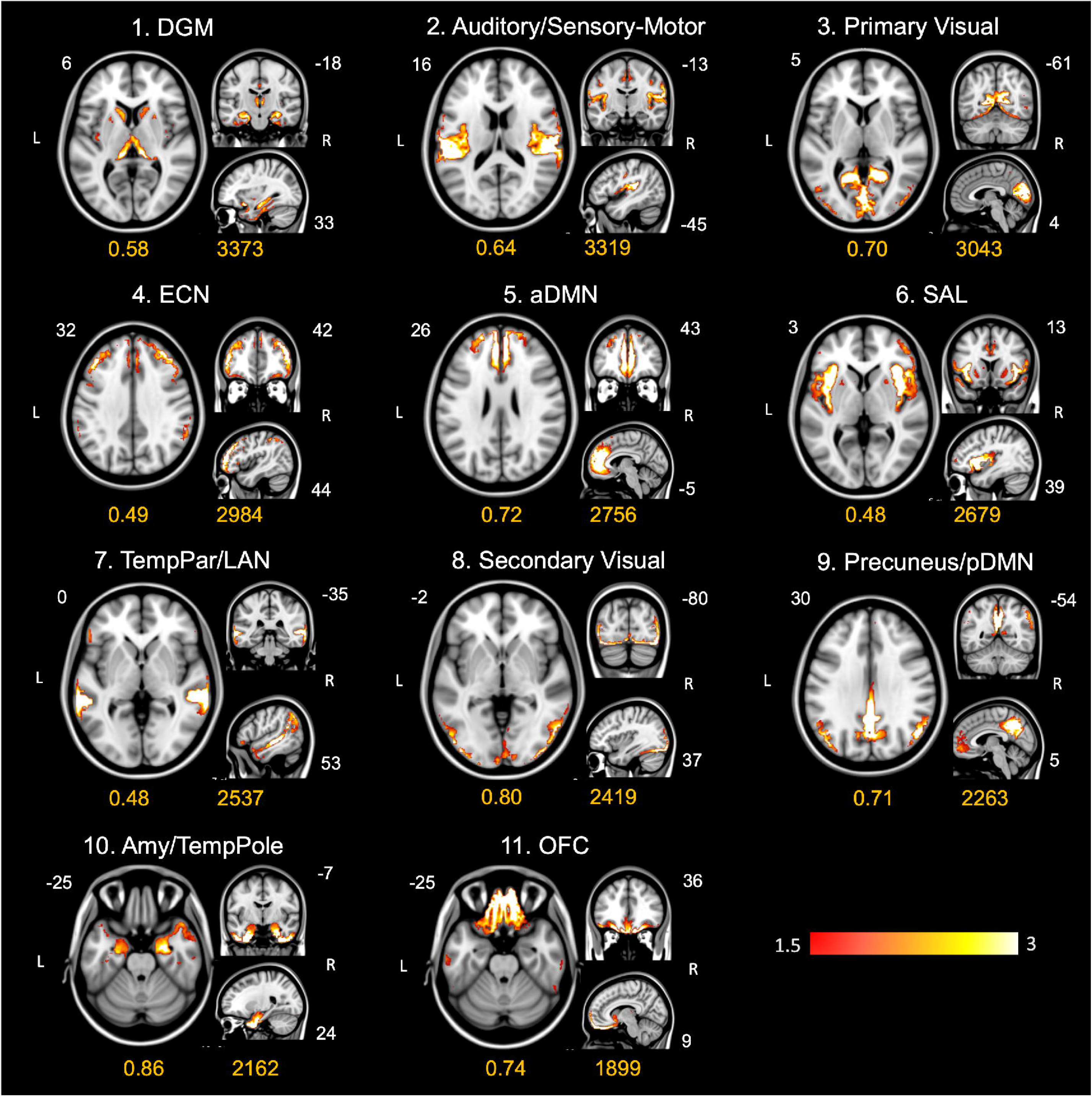
Spatial patterns of the 11 iCAPs retrieved from the analysis on HC and patients with PMS. Under each iCAP are reported the average consensus, and the number of innovation frames assigned to it. Coordinates refer to the Montreal Neurological Institute space. DGM: deep grey matter; Aud/SM: the auditory/sensory-motor; PrimVis: primary visual; ECN: executive control network; aDMN: anterior DMN; SAL: salience; TempPar/Lan: temporo-parietal/language; SecVis: secondary visual; pCun/pDMN: precuneus/posterior DMN; Amy/TP: amygdala/temporal pole; OFC: orbito-frontal cortex.

### Temporal properties of iCAPs: group differences

#### Baseline

The overall duration of the aDMN was significantly reduced in patients with PMS compared to HC (*F*(1,52)=10.99, p= .002, Figure 3A, Table SI 5). A significantly reduced anti-coupling between aDMN and ECN was also observed in patients with PMS compared to HC (*F*(1,52)=7.2, p=.010, Figure 3B, Table SI 6 and 7). No significant differences were detected when iCAPs temporal metrics were compared between CI and CP patients (Table SI 8, 9 and 10).

**Figure 3.**
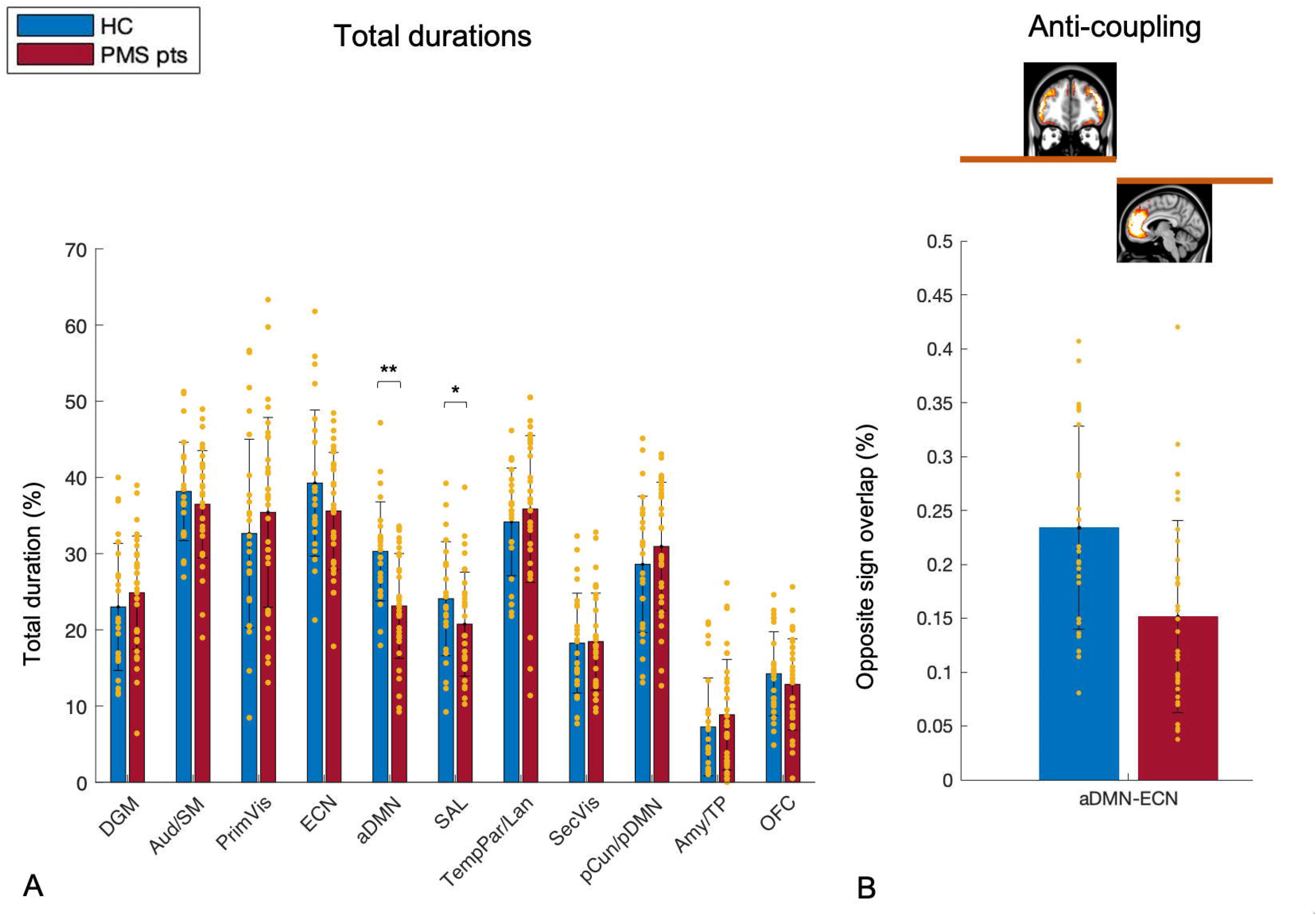
*A*: bar plots of the duration of the 11 iCAPs in HC and patients with PMS. *B*: anti-coupling between ECN and aDMN, resulted significantly different between the HC and PMS patients. DGM: deep grey matter; Aud/SM: the auditory/sensory-motor; PrimVis: primary visual; ECN: executive control network; aDMN: anterior DMN; SAL: salience; TempPar/Lan: temporo-parietal/language; SecVis: secondary visual; pCun/pDMN: precuneus/posterior DMN; Amy/TP: amygdala/temporal pole; OFC: orbito-frontal cortex. *p<0.05, ** surviving FDR correction.

#### FU

The longitudinal analysis did not reveal differences in iCAPs durations in PMS patients or HC at FU vs baseline while the aDMN-ECN coupling significantly increased in PMS patients at FU compared to baseline (*t*(18)=-3.86, *p*= .001, Figure SI 4). No significant group x time interaction was found for the 11 networks durations or for the couplings/anti-couplings involving the aDMN. A group effect was confirmed at FU for aDMN duration (*F*(1,30)=14.5, *p*= .001) and the anti-coupling between aDMN and ECN (*F*(1,30)=12.0, *p*= .002). Meanwhile, a group effect emerged for Aud/SM (*F*(1,30)=13.3, *p*= .001) and the aDMN-Aud/SM anti-coupling (*F*(1,30)=4.2, *p*= .049).

### Temporal properties of iCAPs and structural MRI metrics

No significant associations were found between aDMN duration at baseline and T2 or T1 LV. The aDMN-pCun/pDMN anti-coupling and the coupling between aDMN and primVis were positively correlated to T1 LV and T2 LV (*r*(27)= .40, *p*= .04) (*r*(27)= .41, *p*= .03, respectively). No correlations were found between aDMN temporal metrics and volumetric parameters.

### iCAPs temporal properties and clinical parameters at baseline

#### aDMN duration

No significant results were found for EDSS or motor scores. Cognitive scores were associated to aDMN durations (p=0.030, Figure 4). Specifically, a negative correlation was observed for the SDMT and BVMT-R. No significant results emerged from the grouped PLSC analysis (CI and CP patients considered separately).

**Figure 4.**
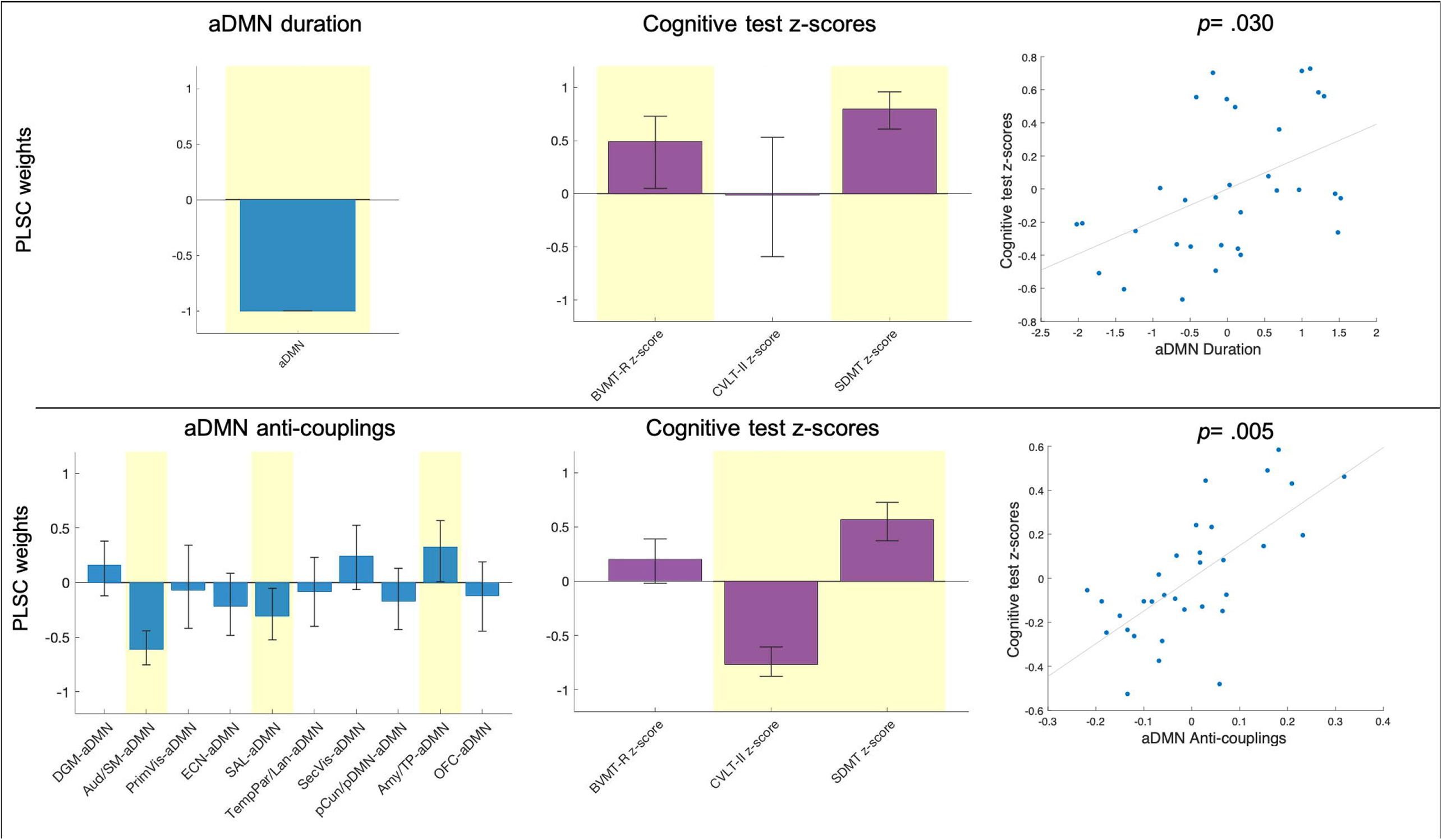
Partial least squares correlation analyses performed to assess the relationship between aDMN duration (top) or anti-couplings (bottom) and cognitive test Z scores. In yellow are shown the robust weights after bootstrapping, and the error bars represent bootstrap intervals. DGM: deep grey matter; Aud/SM: the auditory/sensory-motor; PrimVis: primary visual; ECN: executive control network; aDMN: anterior DMN; SAL: salience; TempPar/Lan: temporo-parietal/language; SecVis: secondary visual; pCun/pDMN: precuneus/posterior DMN; Amy/TP: amygdala/temporal pole; OFC: orbito-frontal cortex; BVMT-R: brief visual memory test revised; CVLT-II: California verbal learning test (second edition); SDMT: symbol digit modalities test.

#### aDMN anti-couplings

A significant association between aDMN anti-couplings and cognitive performance was found (p=0.005, Figure 4) in the whole patients group. In particular, SDMT and CVLT-II were related to aDMN-Aud/SM, aDMN-Amy/TempP, and aDMN-SAL, with the result mainly driven by positive correlations between aDMN-Aud/SM or aDMN-SAL and CVLT-II score and the negative correlations between aDMN-Aud/SM and SDMT and between aDMN-Amy/TempP and CVLT-II. When the grouped PLSC was performed, separating CI and CP patients, we found that aDMN anti-couplings were related to cognitive performance in the CI, but not in the CP group (p= .019, Figure SI 5).

#### All iCAPs durations

This exploratory analysis additionally revealed that the durations of DGM, Aud/SM, TempPar/LAN and pCun/pDMN were significantly correlated to EDSS and T25FW (*p*= .027, Figure SI 6).

### Reproducibility analyses

Analyses to verify the reproducibility of the results on study sub-samples are reported in the SI.

## Discussion

In this work, we characterized the changes in the dynamics of brain functional patterns occurring in patients with PMS patients, showing that they present changes in the functional activation of the aDMN, and in its interplay with ECN. Such changes were detected also over one year FU, in a sample of patients who did not show a significant worsening in clinical disability. Moreover, the dynamic temporal characteristics of the aDMN, namely its reduced sustained activity and the interaction with Aud/SM, SAL and Amy/TempPole, explained cognitive disability in patients with PMS.

Regarding the methodology, we chose to apply this specific data-driven approach since allows to overcome the limitation of the temporal and spatial segregation of functional networks, which characterizes other techniques utilized to investigate functional dynamics, thus better grasping the dynamic interplay between networks over the time of a rs-fMRI sequence. Moreover, these spatial patterns of functional activity have been showed to be reliable across subjects^29^.

As first contribution, this study revealed that the aDMN overall activity duration and its anti-correlation with ECN activity were reduced in patients with PMS. Previous studies showed a reduced aDMN conventional functional activity, related to cognitive disability, in PMS^3^, and highlighted its functional changes in other neurodegenerative disorders^30^. The ECN and DMN are usually oppositely engaged and involved in the switching between the “idling” vs “task-oriented” condition. Besides, together with the SAL, they have been associated to clinical phenotypes in neurodegenerative and psychiatric conditions. Here, we confirm the central role of aDMN dysfunction in the progressive phase of MS and advance the hypothesis an unbalanced and less dynamic interaction with ECN could be linked to the underlying neurodegenerative process. Intriguingly, the anti-coupling between aDMN and pCun/pDMN as well as its coupling with the primVis network were correlated to lesion load, endorsing the connection between structural damage and functional disruption.

A second relevant finding was the absence of significant changes in functional dynamics over time, accompanied by the lack of clinical worsening. Although we cannot exclude this result could be impacted by attrition bias or learning effect for cognitive tests scores, previous studies in PMS propend for slow changes in clinical and structural imaging metrics^31,32^ and the longitudinal variation of brain functional activity might be as gradual. Nonetheless, further longitudinal studies with larger sample sizes are needed to address whether more subtle brain functional modifications occur in the late progressive phase.

As last result, we found a strong relationship between altered functional dynamics of the aDMN and cognitive disability. The DMN plays a pivotal role in the cognitive disability in patients with MS^3,6,33^ and previous studies on the dynamic rs-FC in patients with MS highlighted its role in explaining cognitive impairment, including processing speed^11,13^. Our results indicated that an increased engagement of the aDMN was associated to a worse cognitive performance. Besides, the cognitive domains expressed by the three tests were differently explained by the interaction between aDMN and the other networks. Namely, a better performance in verbal memory, the cognitive domain less affected in our patients’ population, was associated to an increased anti-coupling between aDMN and both Aud/SM and the SAL networks, suggesting the synchronous opposite-signed activation contributes to ensure a better cognitive functioning, while the opposite was observed for the interaction between aDMN and Amy/TempP. Besides, the aDMN-Aud/SM reduced segregation correlated with a better processing speed, suggesting a compensatory role for the aDMN, likely involving its synchronicity with sensory-motor regions^34^. Data from literature suggests an influence of functional dynamics during rest on the subsequent performance during tasks^35^, hence a possible interpretation of the reduced aDMN dynamics as compensatory, in an attempt to adapt to cognitive tasks demand. Another interpretation entails the aDMN interplay with other networks and the reduced overall duration as an attempt to respond to structural damage reducing the dynamic fluidity to privilege efficiency. When a grouped analysis was performed, the aDMN anti-couplings explained the cognitive status in the CI patients, supporting the idea that changes in behavioral performance are better seized by the dynamic interaction among functional networks. In particular, it emerged an additional relationship between a reduced aDMN-pCun/pDMN anti-coupling and better visuo-spatial memory and processing speed. The latter also correlated to a reduced opposite-signed activity of aDMN with ECN and increased anti-correlation with the higher visual network.

An additional exploratory analysis showed a relationship between T25FW and EDSS and the Aud/SM, the TempPar, DGM and pCun/pDMN activity duration. Although none of these networks temporal properties significantly differed, compared to HC, their relative changes in functional engagement might underlie the behavioral modifications characterizing disability in progressive MS.

This study comes with several limitations. First, the small sample size and we cannot exclude type II errors. Second, the high rate of drops-out, which could have biased the results and determined the lack of significant changes we observe over one-year FU. Third, learning effects could have affected the longitudinal results, thus the absence of a significant worsening in cognitive tests at FU compared to baseline. Fourth, motor and cognitive test scores were not available for the HC group, which urges caution in the interpretation of the correlation analyses. Fifth, the use of a multi-band acceleration factor of 7 that could have introduced noise in the rs-fMRI data. Sixth, the lack of an external validation, although the analyses on the sub-samples of our study population proved stable and reproducible results. Seventh, the absence of a population of patients at the earlier stages of the disease, which prevents us to conclude that our findings are specific of the progressive stages of MS.

In conclusion, this study characterized functional brain dynamics in patients with PMS, revealing a reduced engagement of the aDMN *state*, though not detrimental for the cognitive status which is additionally related to the dynamic interplay between aDMN and other functional networks. Future studies are needed to confirm these findings and to address the coupling between functional modifications and local or diffuse structural damage through the different disease stages.

## Data Availability

Data supporting the findings of this study will be shared upon request.

## Abbreviations

PMS: progressive multiple sclerosis
PP: primary progressive
SP: secondary progressive
fMRI: functional MRI
rs-fMRI: resting state functional MRI
DMN: default mode network
rs-FC: resting state functional connectivity
iCAP: innovation driven co-activation pattern
BOLD: blood-oxygenated-level dependent
FU: follow-up
HC: healthy control
EDSS: expanded disability status scale
T25FW: timed 25-foot walking
9HPT: 9-hole peg test
SDMT: symbol digit modalities test
CVLT-II: California verbal learning test second edition
BVMT-R: brief visuospatial memory test revised
WM: white matter
CSF: cerebrospinal fluid
FD: framewise displacement
TA: total activation
MNI: Montreal Neurological Institute
ANCOVA: analysis of covariance
FDR: false discovery rate
PLSC: partial least squares correlation
DGM: deep grey matter
Aud/SM: the auditory/sensory-motor
PrimVis: primary visual
ECN: executive control network
aDMN: anterior DMN
SAL: salience
TempPar/Lan: temporo-parietal/language
SecVis: secondary visual
pCun/pDMN: precuneus/posterior DMN
Amy/TP: amygdala/temporal pole
OFC: orbito-frontal cortex.

