## Supplementary Information for "Altered anterior default mode network dynamics in progressive multiple sclerosis"

### 1 Supplementary Methods

##### Inclusion and exclusion criteria for participants

Diagnosis of PP or SP MS was made according to the revised McDonald's criteria 2010 [1]. Inclusion criteria were the following: i) age between 25 and 65 years, ii) expanded disability status scale (EDSS) score below 6.5 at screening, iii) if treated, patients had to be on a stable treatment for the previous year, iv) no history of relapses in the previous year. Exclusion criteria included: current or past history of major hematological, renal, hepatic, psychiatric or neurological disease (other than MS for the patient group) and contraindications to MRI. From the initial sample of 74 subjects, 4 patients were excluded because of failure in MRI processing (i.e. functional-structural co-registration) and 13 (1 healthy control, 12 patients) were excluded after motion scrubbing because more than 25% of volumes in their scans had a framewise displacement above the threshold of 0.5 mm (see "Motion" section), resulting in a final sample of 57 subjects.

##### MRI sequences parameters

T2-weighted sequence: voxel size 0.5x0.5x3 mm, repetition time (TR) 8000 ms, echo time (TE) 95ms. 3D T1-weighted sequence: voxel size:0.8x0.8x0.8 mm, TR 3000 ms, TE 2.47 ms, flip angle 7°, inversion time 1000 ms). Single shot gradient echo planar imaging sequence: voxel size: 2.1x2.1x2.1 mm, TR 1000 ms, TE 35 ms, multi-band acceleration factor 7, flip angle 60°, 400 volumes. During rs-fMRI, subjects were asked to rest with their eyes closed.

##### Motion

We identified subjects that displayed too much motion characterized by their framewise displacement values [2]. Those with FDPower $> 0.5$ mm in more than 25% of the volumes [3] were excluded from the analysis. In the final sample, the average FD did not differ between patients with PMS and HC at the baseline (t(55)= -1.37, p=0.18) and at FU (t(33)= -1.52, p=0.14, see Table 1). In all other subjects, scrubbing was applied with contaminated volumes (FDPower $> 0.5$mm) censored and replaced by the spline interpolation of the previous and following volumes. Motion frames were excluded from the computation of iCAPs temporal properties.

##### Total activation and iCAPs extraction

The iCAPs analysis was performed using a publicly available implementation (`https://c4science.ch/source/iCAPs/`). First, a subsequent denoising and hemodynamic deconvolution was applied using total activation (TA) to the single subject's native space rs-fMRI preprocessed time-courses, both at baseline and FU. This method allows to analyze the fMRI signal independently from the hemodynamic response, employing the changes of signal induced by neuronal activity. [4]. For each voxel $v$ at each moment $t$, the time course results as $\mathbf{y}[v,t] = \mathbf{x}[v,t] + \varepsilon$, where the BOLD activity-related signal $\mathbf{x}$ can bee described by $x[v,t] = \Sigma_{\tau=0}^{t}\mathbf{s}[v,t]\mathbf{h}(t-\tau)$, and $\mathbf{s}$ describes the changes induced by neural activity. The activity related signal **x** can be retrieved through a spatio-temporal regularization as follows:

$$\hat{\mathbf{x}} = \underset{x}{\operatorname{argmin}} \left\{ \frac{1}{2} \|\mathbf{y} - \mathbf{x}\|_2^2 + \mathcal{R}_T(\mathbf{x}) + \mathcal{R}_S(\mathbf{x}) \right\}$$

.

The temporal regularization can be expressed as:

$$\mathcal{R}_T(\mathbf{x}) = \sum_{v=1}^{N_x} \lambda_T(v) \sum_{t=1}^{N_t} |\Delta_L \mathbf{x}[v,t]|$$

where $\Delta_L$ is the discrete version of the differential operator $L$ and $\lambda_T(v)$ is the regularization parameter for voxel $(v)$.

The spatial regularization is given by:

$$\mathcal{R}_S(\mathbf{x}) = \sum_{t=1}^{N_t} \lambda_S(t) \sum_{v=1}^{N_x} \left( \sum_{u \in \mathcal{N}(v)} \left(\mathbf{x}[v,t] - \mathbf{x}[u,t]\right)^2 \right)^{1/2}$$

where $\mathcal{N}(v)$ is a set of neighor voxels around $v$ and $\lambda_S(v)$ is the spatial regularization parameter at a given timepoint $t$. For more details about the implementation of the TA paradigm we refer to [5] and [4].

After running TA, the sparse innovation signal (or transients) are extracted by computing the temporal derivative of the activity-inducing signals. Afterwards, the significant transients are extracted by taking only a fraction across brain volumes following a two-step thresholding procedure. The temporal thresholding includes a phase-randomization, from which the cut-off thresholds using the lowest 5th and highest 95th percentile of its corresponding innovation signal are extracted. Following the temporal thresholding, the frames with at least $5\%$ of the total number of gray matter voxels are selected to undergo temporal clustering using k-means. For more details of the iCAPs extraction and the optimization of the thresholding, we refer to [6]. Thereafter, rs-fMRI frames corresponding to transients were normalized to the MNI coordinate space and concatenated across all subjects. The resulting matrix of dimension "number of voxels x transients" underwent temporal k-means clustering, to obtain brain patterns that are simultaneously transitioning, i.e. iCAPs. Consensus clustering was used to determine the optimum number of clusters. The time-courses "assigned" to each iCAP were Z-scored within each subject and thresholded at $|z| > 1$ to obtain binary data and be able to define the "active" time-points. The choice of threshold was motivated by previous works that implemented TA and iCAPs framework [6]–[8]. Lastly, the activity inducing time-courses for each subject were obtained for all iCAPs using spatio-temporal transient-informed regression[7]. ICAPs duration is defined as the percentage of the active time-points with respect to the total time-points for each iCAP. Couplings and anti-couplings for each pair of iCAPs were derived using the Jaccard index and defined as follow: i) the coupling: number of time-points during which the two iCAPs were both co-(de)active divided by the number of time points during which at least one of them was (de)active, and ii) the anti-coupling: number of time-points during which the two iCAPs presented opposite sign of activation, divided by the number of time points during which at least one of them was (de)active. For the computation of duration and couplings, only activation blocks of two or more time-points have been considered.

**Choice of the optimal number of clusters**

Consensus clustering [9] was used to find the optimal number of clusters in the concatenated significant innovation frames. This method includes a subsampling of the data and multiple runs of the clustering algorithm. The consistency of each frame to be grouped in a similar cluster is monitored through a consensus metric. The consensus clustering matrices for each evaluated K are represented in Figure SI1 for the baseline sample. The distribution of each cluster consensus is plotted in Figure SI2. Based on the highest median consensus we chose a K= 13 [9]. ICAPs determined by less than 5 subjects were discarded from further analysis.

Figure SI 1: Consensus clustering matrices for different cluster values K = 11 to 21 for baseline (57 subjects) data. Axes represents frames number. Values in the matrix range from 0 to 1, indicating the reproducibility of the sampling across multiple runs, with 1 being perfectly re-sampled at all times. Diagonal values are expected to be equal to 1 (the same frame indices will always be clustered into the same group).

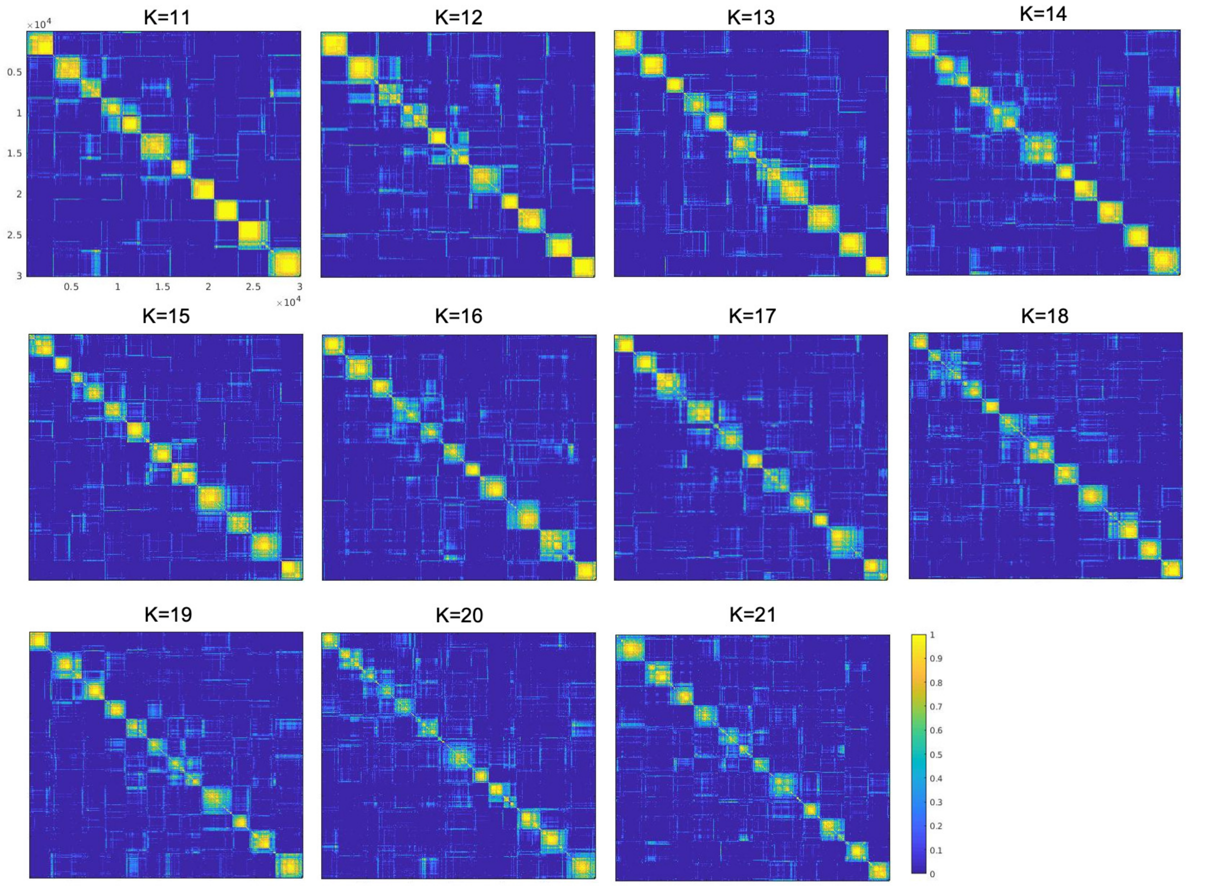

Figure SI 2: Clustering consensus as a measure of the stability of the observed iCAPs with respect to different K (x axis) over multiple runs of the clustering

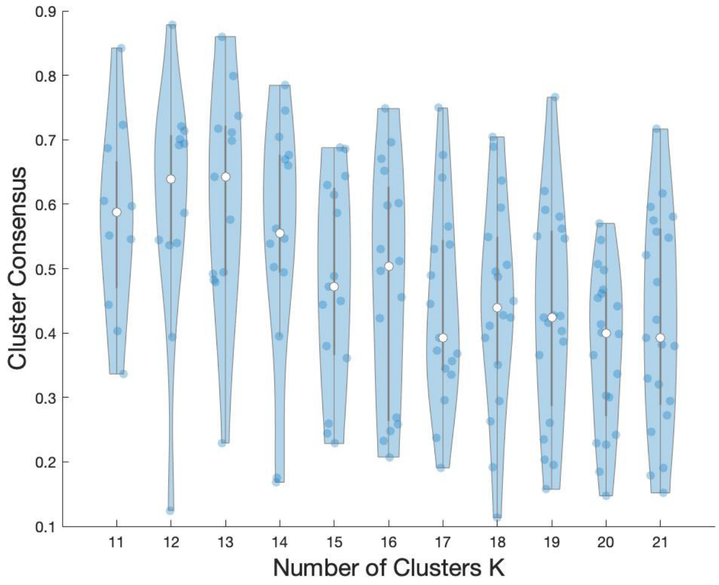


#### Analysis at follow-up

The TA analysis was applied to the 35 subjects at follow-up (FU). Then, the iCAPs retrieved from the baseline sample were fitted into the FU sample. This was done by matching each significant innovation frame of subjects at FU to the closest iCAP obtained from baseline using cosine distance, as a metric. After obtaining the cluster index for the innovation frames at FU and their distance to each cluster center, the transient-informed regression step was repeated on FU data to obtain each subject's iCAP time-courses and their corresponding temporal properties.

#### Partial least squares correlation analysis

Partial least squares correlation (PLSC) analysis [10] procedure included: first, the [subjects x temporal properties] matrix X for brain variables (iCAPs temporal properties) and the [subjects x behavioral variables] matrix Y for clinical ones were built, concatenating HC and PMS patients. Then, their relationship was assessed by means of singular value decomposition of the correlation matrix between X and Y, which resulted in latent components that indicate multivariate patterns of brain-behavior correlation. Lastly, the significance and stability of the components was assessed by permutation (1000 permutations) and bootstrap (800 bootstrap samples) testing, respectively.

### 2 Supplementary Results

**Contribution of each group to the number of frames selected for each iCAP**

To evaluate whether the 2 groups (HC and MS pts) significantly differed in the contribution of frames to each iCAP, we retrieved, for each subject, the number of frames with which they contributed to each iCAP and compared the two groups using the Wilcoxon rank sum test, given the non normal distribution of the data. All p values resulted ¿ 0.05. Mean, median and SD values for number of frames with which HC or MS pts groups contributed to each iCAP are reported in the Table SI 4.

**Frequency of cognitive impairment**

Twenty-four (75%), 13 (68%) and 8 (42%) patients were classified as CI at baseline, in the sub-sample (BS-Sub) of 35 patients at baseline and at FU, respectively. A chi-squared test did not find significant differences in the number of CI patients between the BS-Sub and FU groups. Performance at BS-Sub and FU timepoints were significantly correlated for the three tests (Pearson correlation coefficients range: 0.46-0.56). (Figure SI 3).

Table SI 1: Clinical and demographic data at baseline of the drop-out subjects.

| | Drop-out subjects (n=22) | | | |
|---|---|---|---|---|
| | HC (n=9) | p-value‡ | PMS patients (n=13) | p-value† |
| Age | $46.2 \pm 11.4$ | 0.66 | $49.5 \pm 10.7$ | 0.55 |
| Gender (females, males) | 3; 6 | 0.61 | 7; 6 | 0.40 |
| Education | $15.6 \pm 0.9$ | 0.25 | $16.2 \pm 2.7$ | 0.49 |
| Disease duration | - | - | $17.2 \pm 11.8$ | 0.50 |
| EDSS (median; range) | - | - | 5.5; 3.0-6.0 | 0.82 |
| SDMT raw score | - | - | $42.6 \pm 15.0$ | 0.27 |
| SDMT Z-score | - | - | $-1.8 \pm 1.5$ | - |
| BVMT-R raw score | - | - | $16.8 \pm 7.0$ | 0.92 |
| BVMT-R Z-score | - | - | $-2.0 \pm 1.3$ | - |
| CVLT-II raw score | - | - | $51.1 \pm 15.0$ | 0.19 |
| CVLT-II Z-score | - | - | $-0.19 \pm 1.5$ | - |
| T25FW | - | - | $7.3 \pm 2.4$ | 0.11 |
| 9HPT | - | - | $33.2 \pm 9.1$ | 0.64 |
| T2 LV (mL) | - | - | $10.0 \pm 11.0$ | 0.28 |
| T1 LV (mL) | - | - | $8.0 \pm 7.1$ | 0.22 |
| Grey Matter Volume ($cm^3$) | $790.3 \pm 31.4$ | 0.38 | $747.2 \pm 51.7$ | 0.35 |
| White Matter Volume ($cm^3$) | $720.4 \pm 37.7$ | 0.74 | $655.6 \pm 40.1$ | 0.80 |
| Brain Volume ($cm^3$) | $1510.7 \pm 65.5$ | 0.47 | $1402.9 \pm 86.2$ | 0.69 |
| aDMN duration | $31.8 \pm 5.4$ | 0.39 | $24.8 \pm 7.2$ | 0.28 |
| aDMN-ECN anti-coupling | $0.23 \pm 0.1$ | 0.96 | $0.18 \pm 0.1$ | 0.14 |

All variables are expressed as mean ± standard deviation if not otherwise specified. ‡ unpaired t-test between HC who undergo FU and HC who dropped out. † unpaired t-test between PMS patients who undergo FU and PMS patients who dropped out. EDSS: expanded disability status scale; SDMT: symbol digit modalities test; BVMT-R: Brief Visuospatial Memory Test–Revised; CVLT-II: CVLT-II California Verbal Learning Test Second Edition; T25FW: timed 25-foot walk test; 9HPT: 9-hole peg test; T2 LV: lesion volume in T2-weighted sequence; T1 LV: lesion volume in T1-weighted sequence.

Table SI 2: Clinical data of the subsample of subjects who underwent the follow-up analysis at baseline (BS-Sub) and Follow-up.

| | BS-Sub (n=19) | Follow-up (n= 19) | p-value‡ |
|---|---|---|---|
| EDSS (median; range) | 6; 1.0-6.5 | 5; 2.5-6.5 | 0.59 |
| SDMT raw score | $48.8 \pm 15.7$ | $51.3 \pm 13.3$ | 0.28 |
| BVMT-R raw score | $16.5 \pm 9.9$ | $19.8 \pm 8.9$ | 0.06 |
| CVLT-II raw score | $57.3 \pm 11.6$ | $58.7 \pm 13.3$ | 0.38 |
| T25FW | $9.8 \pm 5.2$ | $12.1 \pm 15.8$ | 0.47 |
| 9HPT | $31.4 \pm 11.4$ | $34.2 \pm 20.3$ | 0.41 |
| T2 LV (mL) | $6.8 \pm 9.8$ | $7.0 \pm 10.0$ | 0.13 |
| T1 LV (mL) | $4.7 \pm 6.9$ | $5.5 \pm 8.1$ | 0.02 |

All variables are expressed as mean ± standard deviation if not otherwise specified. ‡ paired t-test between BS-Sub and FU PMS patients. BS-Sub: subsample of 35 subjects at baseline; EDSS: expanded disability status scale; SDMT: symbol digit modalities test; BVMT-R: Brief Visuospatial Memory Test–Revised; CVLT-II: CVLT-II California Verbal Learning Test Second Edition; T25FW: timed 25-foot walk test; 9HPT: 9-hole peg test; T1 LV: lesion volume in T1-weighted sequence; T2 LV: lesion volume in T2-weighted sequence.

**Validation analyses**

To evaluate the robustness and reliability of our results, we performed three validation analyses, performing the k-means clustering using the optimal k=13 (as resulted from the Consensus Clustering) and the transient-informed regression steps:

1. on the baseline sample of 57 subjects to assess the robustness of the Clustering step. The same iCAPs as in the original analysis were retrieved. Among iCAPs durations, the aDMN duration resulted significantly reduced in patients with PMS, compared to HC (F(1,52)=12.00, p= .001).

2. on the baseline sub-sample of 35 subjects (16 HC and 19 patients with PMS) who underwent FU. Nine out of the 11 iCAPs were retrieved from these analysis (the last 4 were discarded as determined by less than 5 subjects) and represented in Figure SI 8. Among iCAPs durations, the aDMN duration resulted significantly different between HC and patients with PMS (F(1,30)=6.77, p= .014, reduced in PMS patients).

3. on a sub-sample of 40 subjects, balancing the number of HC (n=20) and patients with PMS (n=20). Ten out of the 11 iCAPs were retrieved from these analysis (the last 3 were discarded as determined by less than 5 subjects). Among iCAPs durations, the aDMN duration resulted significantly reduced in patients with PMS compared to HC (F(1,35)=15.50, p¡ .001).

Figure SI 3: Number of patients performing below the cut-off scores of 1.5 SD using the regression-based norms as in [11] at baseline (BS-Sub, n=19) and at the 1 year follow-up (n=19), and correlations between the test performances at the two time points.

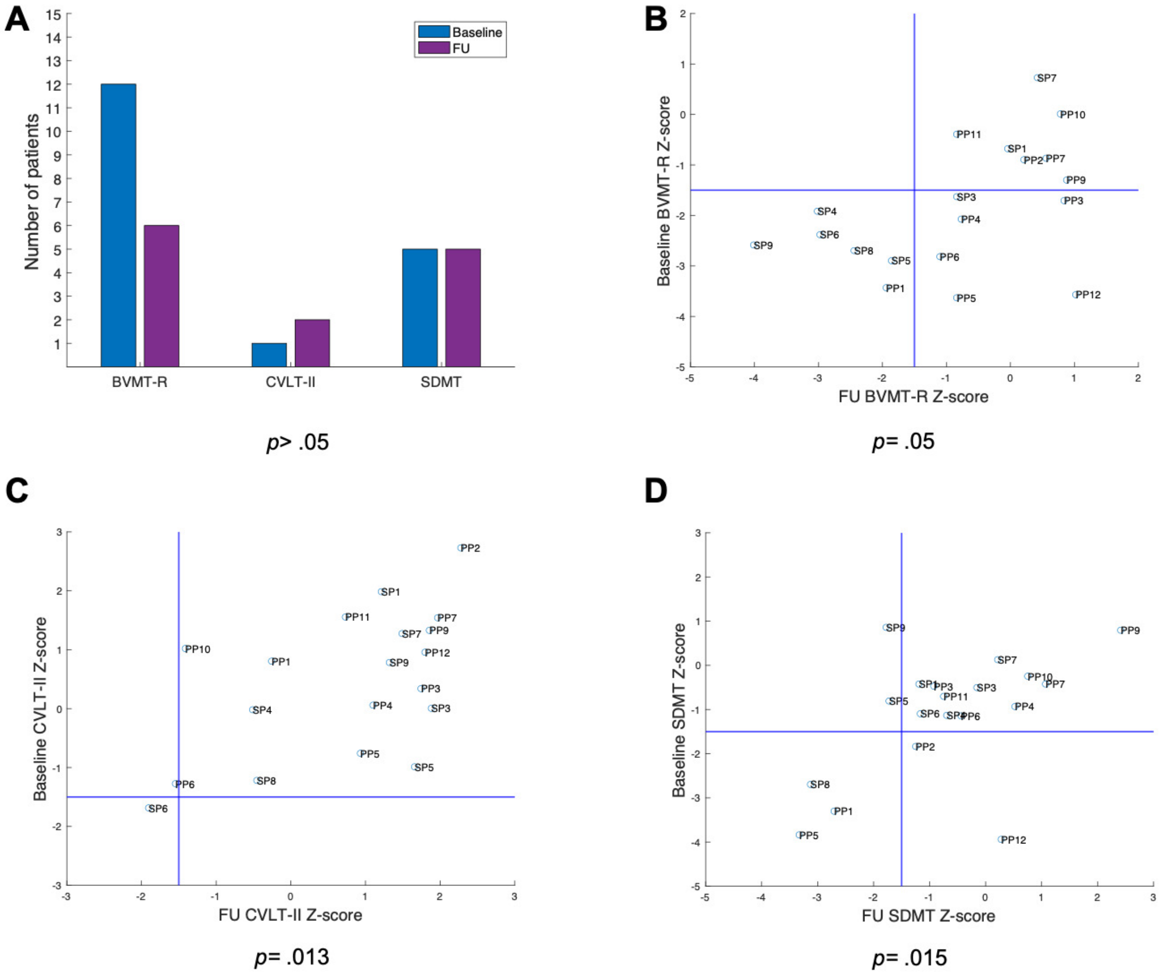

Table SI 3: Baseline iCAPs corresponding to the 17 networks of Yeo atlas [12], the Greicius atlas [13] and the regions in the automated anatomical labeling atlas ([14]). Percentiles indicate the fraction of voxels belonging to a network or region that has a z-score $> 1.5$. Only networks and regions that have at least 25% surviving percentile are included in the list.

| iCAP | Network Schaefer(%) | Network Shirer (%) | Lobe | Region AAL (%) | mean z-score | voxels |
|---|---|---|---|---|---|---|
| 1 | DefaultC (43,12) | Basal Ganglia (56,31) | Limbic | Amygdala L (91,57) | 3,03 | 152 |
| | Limbic2 (31,46) | | Limbic | Amygdala R (86,3) | 3,07 | 126 |
| | | | Limbic | Hippocampus L (86,14) | 2,45 | 491 |
| | | | Limbic | Hippocampus R (84,05) | 2,66 | 432 |
| | | | Subcortical | Thalamus R (80,42) | 2,65 | 193 |
| | | | Subcortical | Caudate L (73,7) | 2,12 | 339 |
| | | | Subcortical | Caudate R (72,85) | 2,14 | 314 |
| | | | Subcortical | Thalamus L (72,82) | 2,48 | 217 |
| | | | Subcortical | Putamen L (71,82) | 2,39 | 209 |
| | | | Limbic | ParaHippocampal L (70,28) | 2,51 | 395 |
| | | | Subcortical | Putamen R (63,4) | 2,42 | 220 |
| | | | Limbic | ParaHippocampal R (61,8) | 2,51 | 419 |
| | | | Frontal | Olfactory R (54,84) | 1,96 | 85 |
| | | | Frontal | Olfactory L (49,73) | 2,02 | 92 |
| | | | Occipital | Fusiform L (41,9) | 2,09 | 538 |
| | | | Temporal | Temporal Pole Sup L (35,5) | 2,28 | 197 |
| | | | Temporal | Temporal Pole Sup R (34,88) | 2,22 | 195 |
| | | | Occipital | Fusiform R (32,58) | 1,98 | 463 |
| 2 | SomMotB (77,87) | Auditory (89,05) | Central | Rolandic Oper L (89,44) | 2,56 | 466 |
| | SalVentAttnA (53,05) | post Salience (42,88) | Central | Rolandic Oper R (89,19) | 2,87 | 561 |
| | SomMotA (27,66) | | Temporal | Heschl R (85,55) | 2,42 | 148 |
| | | | Temporal | Heschl L (74,65) | 2,24 | 106 |
| | | | Parietal | SupraMarginal L (67,71) | 2,68 | 239 |
| | | | Temporal | Temporal Sup L (66,25) | 2,65 | 642 |
| | | | Parietal | SupraMarginal R (55,09) | 2,88 | 341 |
| | | | Temporal | Temporal Sup R (52,31) | 2,68 | 633 |
| | | | Limbic | Insula R (50) | 2,14 | 543 |
| | | | Parietal | Postcentral R (46,78) | 2,34 | 269 |
| | | | Parietal | Postcentral L (44,82) | 2,36 | 277 |
| | | | Limbic | Insula L (42,61) | 2,13 | 548 |
| | | | Frontal | Supp Motor Area R (37,58) | 2,03 | 177 |
| | | | Frontal | Frontal Inf Oper R (36,73) | 1,94 | 119 |
| | | | Limbic | Cingulum Mid L (32,88) | 1,96 | 316 |
| | | | Limbic | Cingulum Mid R (30,88) | 1,98 | 348 |
| | | | Frontal | Precentral R (30,41) | 2,03 | 111 |
| | | | Frontal | Supp Motor Area L (29,03) | 2,02 | 126 |
| 3 | VisPeri (87,77) | prim Visual (97,41) | Occipital | Cuneus R (95,62) | 2,69 | 371 |
| | VisCent (60,94) | high Visual (69,93) | Occipital | Calcarine R (94,5) | 3,04 | 687 |
| | DefaultC (28,1) | | Occipital | Cuneus L (91,34) | 2,55 | 369 |
| | | | Occipital | Calcarine L (87,44) | 2,78 | 787 |
| | | | Occipital | Occipital Sup L (86,42) | 2,43 | 140 |
| | | | Occipital | Occipital Mid R (80,69) | 2,42 | 418 |
| | | | Occipital | Occipital Sup R (78,62) | 2,18 | 125 |
| | | | Occipital | Occipital Mid L (69,3) | 2,33 | 526 |
| | | | Occipital | Lingual L (65,19) | 2,54 | 472 |
| | | | Occipital | Lingual R (64,42) | 2,62 | 603 |
| | | | Occipital | Occipital Inf L (41,67) | 1,83 | 105 |
| | | | Occipital | Fusiform R (29,56) | 1,88 | 420 |

| iCAP | Network Schaefer (%) | Network Shirer (%) | Lobe | Region AAL (%) | mean z-score | voxels |
|---|---|---|---|---|---|---|
| 4 | ContA (70,45) | RECN (83,27) | Frontal | Frontal Mid R (87,95) | 2,85 | 1212 |
| | SalVentAttnB (53,83) | antSalience (45,45) | Parietal | Parietal Inf R (78,72) | 2,48 | 270 |
| | ContB (46,41) | LECN (40,02) | Frontal | Frontal Inf Tri R (76,47) | 2,44 | 390 |
| | | Visuospatial (33,2) | Frontal | Frontal Mid L (73,99) | 2,55 | 879 |
| | | post Salience (30,77) | Frontal | Frontal Mid Orb R (66,19) | 2,44 | 327 |
| | | | Frontal | Frontal Inf Tri L (58,52) | 2,46 | 364 |
| | | | Frontal | Frontal Sup R (55,87) | 2,27 | 414 |
| | | | Frontal | Frontal Mid Orb L (51,27) | 2,45 | 222 |
| | | | Parietal | Parietal Inf L (46,61) | 2,05 | 213 |
| | | | Frontal | Frontal Inf Oper R (41,) | 1,9 | 136 |
| | | | Frontal | Frontal Sup L (39,85) | 1,99 | 263 |
| | | | Parietal | SupraMarginal R (38,45) | 2,34 | 238 |
| | | | Frontal | Frontal Sup Medial R (34,72) | 2,21 | 251 |
| | | | Frontal | Frontal Inf Oper L (34,58) | 1,85 | 102 |
| | | | Frontal | Frontal Sup Medial L (25,57) | 2,1 | 212 |
| 5 | DefaultA (47,3) | dorsalDMN (65,09) | Limbic | Cingulum Ant R (89,64) | 3,45 | 554 |
| | SalVentAttnB (44,43) | antSalience (34,36) | Frontal | Frontal Sup Medial R (89,63) | 2,82 | 648 |
| | | | Frontal | Frontal Sup Medial L (89,02) | 3,15 | 738 |
| | | | Limbic | Cingulum Ant L (85,54) | 3,46 | 704 |
| | | | Frontal | Frontal Sup Orb Medial R (81,63) | 2,64 | 360 |
| | | | Frontal | Frontal Sup Orb Medial L (79,14) | 2,57 | 330 |
| | | | Frontal | Frontal Sup L (64,7) | 2,15 | 427 |
| | | | Frontal | Frontal Sup R (34,82) | 1,98 | 258 |
| | | | Frontal | Rectus L (26,77) | 2 | 140 |
| 6 | SalVentAttnA (50,53) | antSalience (42,34) | Limbic | Insula L (87,33) | 2,82 | 1123 |
| | SomMotB (27,47) | Auditory (33,06) | Limbic | Insula R (85,36) | 2,94 | 927 |
| | SalVentAttnB (25,23) | | Frontal | Frontal Inf Oper R (54,63) | 2,43 | 177 |
| | | | Frontal | Frontal Inf Tri R (53,33) | 2,24 | 272 |
| | | | Subcortical | Putamen R (52,45) | 1,76 | 182 |
| | | | Subcortical | Putamen L (51,89) | 1,79 | 151 |
| | | | Central | Rolandic Oper L (48,56) | 2,03 | 253 |
| | | | Frontal | Frontal Inf Oper L (46,78) | 2,01 | 138 |
| | | | Temporal | Heschl L (42,25) | 1,81 | 60 |
| | | | Central | Rolandic Oper R (37,68) | 2,05 | 237 |
| | | | Frontal | Frontal Inf Orb R (33,15) | 2,17 | 241 |
| | | | Temporal | Temporal Pole Sup L (31,71) | 1,99 | 176 |
| | | | Limbic | Cingulum Ant R (31,07) | 1,82 | 192 |
| | | | Frontal | Frontal Inf Tri L (28,94) | 2,26 | 180 |
| | | | Temporal | Temporal Pole Sup R (28,44) | 1,91 | 159 |
| 7 | TempPar (75,35) | Language (78,83) | Parietal | Angular L (81,14) | 2,63 | 241 |
| | DefaultB (38,16) | | Temporal | Temporal Mid L (76,58) | 2,82 | 1383 |
| | | | Temporal | Temporal Mid R (67,69) | 2,84 | 1280 |
| | | | Parietal | Angular R (55,02) | 2,51 | 252 |
| | | | Temporal | Temporal Sup R (39,01) | 2,66 | 472 |
| | | | Temporal | Temporal Pole Sup L (30,63) | 1,98 | 170 |
| | | | Parietal | Parietal Inf R (25,07) | 2,36 | 86 |
| 8 | VisCent (67,65) | high Visual (82,47) | Occipital | Occipital Inf R (98,47) | 3,59 | 258 |
| | DorsAttnA (52,55) | | Occipital | Occipital Inf L (91,67) | 3,01 | 231 |
| | | | Temporal | Temporal Inf R (47,23) | 2,82 | 894 |
| | | | Occipital | Occipital Mid R (46,53) | 2,34 | 241 |
| | | | Occipital | Occipital Mid L (45,45) | 2,21 | 345 |
| | | | Occipital | Fusiform R (37,37) | 2,41 | 531 |
| | | | Occipital | Lingual R (32,16) | 2,77 | 301 |
| | | | Temporal | Temporal Inf L (29,87) | 2,55 | 463 |
| | | | Occipital | Fusiform L (29,36) | 2,6 | 377 |
| | | | Occipital | Calcarine L (29) | 2,25 | 261 |

| iCAP | Network Schaefer (%) | Network Shirer (%) | Lobe | Region AAL (%) | mean z-score | voxels |
|---|---|---|---|---|---|---|
| 9 | ContC (70,11) | Precuneus (66,46) | Limbic | Cingulum Post L (99,43) | 4,02 | 174 |
| | DefaultA (50,89) | ventralDMN (48,38) | Limbic | Cingulum Post R (99,07) | 4,25 | 106 |
| | | dorsalDMN (37,96) | Parietal | Angular L (90,57) | 2,47 | 269 |
| | | | Parietal | Angular R (89,52) | 2,75 | 410 |
| | | | Parietal | Precuneus R (73,42) | 3,26 | 721 |
| | | | Parietal | Precuneus L (68,91) | 3,15 | 614 |
| | | | Frontal | Frontal Sup Orb Medial R (42,63) | 1,87 | 188 |
| | | | Frontal | Frontal Sup Orb Medial L (42,45) | 1,85 | 177 |
| | | | Occipital | Cuneus R (30,67) | 2,11 | 119 |
| | | | Limbic | Cingulum Mid R (27,15) | 3,15 | 306 |
| | | | Limbic | Cingulum Mid L (26,74) | 2,82 | 257 |
| | | | Occipital | Cuneus L (26,49) | 2,27 | 107 |
| | | | Parietal | Parietal Inf R (26,24) | 2,29 | 90 |
| 10 | Limbic2 (74,67) | | Temporal | Temporal Pole Mid R (91,36) | 3,11 | 476 |
| | | | Temporal | Temporal Pole Mid L (86,98) | 2,77 | 354 |
| | | | Limbic | Amygdala R (67,81) | 2,08 | 99 |
| | | | Limbic | Amygdala L (63,25) | 1,98 | 105 |
| | | | Temporal | Temporal Pole Sup R (53,67) | 2,09 | 300 |
| | | | Limbic | ParaHippocampal R (47,2) | 2,69 | 320 |
| | | | Limbic | ParaHippocampal L (42,35) | 2,72 | 238 |
| | | | Limbic | Hippocampus R (41,25) | 2,27 | 212 |
| | | | Temporal | Temporal Inf L (36,45) | 3,25 | 565 |
| | | | Limbic | Hippocampus L (32,98) | 1,98 | 188 |
| | | | Temporal | Temporal Inf R (32,17) | 3,32 | 609 |
| 11 | Limbic1 (88,89) | | Frontal | Frontal Sup Orb R (95,1) | 3,25 | 408 |
| | ContB (31,29) | | Frontal | Frontal Sup Orb L (88,67) | 3,18 | 407 |
| | | | Frontal | Rectus L (86,62) | 2,76 | 453 |
| | | | Frontal | Rectus R (82,84) | 2,94 | 367 |
| | | | Frontal | Frontal Mid Orb L (78,98) | 3,24 | 342 |
| | | | Frontal | Frontal Mid Orb R (75,3) | 3,16 | 372 |
| | | | Frontal | Olfactory L (69,19) | 2,19 | 128 |
| | | | Frontal | Olfactory R (68,39) | 2,14 | 106 |
| | | | Frontal | Frontal Sup Orb Medial L (56,35) | 2,23 | 235 |
| | | | Frontal | Frontal Inf Orb L (53,96) | 2,41 | 436 |
| | | | Frontal | Frontal Sup Orb Medial R (49,21) | 2,33 | 217 |
| | | | Subcortical | Caudate L (42,83) | 1,91 | 197 |
| | | | Frontal | Frontal Inf Orb R (36,86) | 2,24 | 268 |
| | | | Subcortical | Caudate R (35,5) | 1,92 | 153 |

Table SI 4: Median, mean and SD of the number of frames each group contributed to each iCAP and p values of the resulting Wilcoxon rank sum test

| | Median HC | Median MS pts | Mean HC | Mean MS pts | SD HC | SD MS pts | p value |
|---|---|---|---|---|---|---|---|
| 1. Deep GM | 47.00 | 57.50 | 57.12 | 60.78 | 31.68 | 36.64 | .60 |
| 2. Auditory/Sensory-Motor | 52.00 | 48.50 | 57.28 | 58.97 | 30.96 | 32.10 | .74 |
| 3. Primary Visual | 48.00 | 51.50 | 51.88 | 54.56 | 21.94 | 21.13 | .53 |
| 4. ECN | 46.00 | 49.50 | 50.00 | 51.38 | 22.60 | 24.19 | .65 |
| 5. aDMN | 45.00 | 43.00 | 51.56 | 45.84 | 23.22 | 20.68 | .43 |
| 6. SAL | 42.00 | 42.50 | 49.36 | 45.16 | 22.77 | 21.86 | .61 |
| 7. TempPar/LAN | 38.00 | 44.00 | 42.08 | 46.41 | 23.32 | 26.66 | .64 |
| 8. Secondary Visual | 41.00 | 40.50 | 42.52 | 42.38 | 23.49 | 22.96 | .92 |
| 9. pCun/pDMN | 36.00 | 35.00 | 36.64 | 42.09 | 17.57 | 20.63 | .49 |
| 10. Amy/TempPole | 19.00 | 31.50 | 34.64 | 40.50 | 33.28 | 31.62 | .45 |
| 11. OFC | 27.00 | 33.00 | 30.24 | 35.72 | 14.70 | 19.34 | .27 |

Table SI 5: Results of the ANCOVA analysis to assess differences in iCAPs duration between HC and patients with PMS. Effect size is reported as Cohen's d.

| iCAP | p value | F(1,52) | effect size (Cohen's d) |
|---|---|---|---|
| 1. Deep GM | 0.49 | 0.48 | -0.24 |
| 2. Auditory/Sensory-Motor | 0.33 | 0.99 | 0.25 |
| 3. Primary Visual | 0.86 | 0.03 | -0.22 |
| 4. ECN | 0.59 | 0.30 | 0.43 |
| 5. aDMN | 0.002 | 10.99 | 1.07 |
| 6. SAL | 0.01 | 6.98 | 0.47 |
| 7. TempPar/LAN | 0.65 | 0.20 | -0.20 |
| 8. Secondary Visual | 0.73 | 0.12 | -0.03 |
| 9. pCun/pDMN | 0.55 | 0.37 | -0.27 |
| 10. Amy/TempPole | 0.67 | 0.19 | -0.23 |
| 11. OFC | 0.36 | 0.85 | 0.24 |

Table SI 6: Results of the ANCOVA analysis to assess differences in iCAPs Couplings between HC and patients with PMS. Effect size is reported as Cohen's d.

| iCAPs pair | p value | F(1,52) | effect size (Cohen's d) |
|---|---|---|---|
| Couplings | | | |
| 1-2 | 0.31 | 1.06 | -0.17 |
| 1-3 | 0.24 | 1.39 | -0.36 |
| 2-3 | 0.06 | 3.77 | 0.46 |
| 1-4 | 0.68 | 0.17 | 0.07 |
| 2-4 | 0.59 | 0.30 | 0.30 |
| 3-4 | 0.98 | 0.00 | -0.16 |
| 1-5 | 0.37 | 0.83 | 0.26 |
| 2-5 | 0.78 | 0.08 | 0.21 |
| 3-5 | 0.09 | 3.07 | 0.53 |
| 4-5 | 0.27 | 1.26 | -0.47 |
| 1-6 | 0.38 | 0.79 | 0.28 |
| 2-6 | 0.10 | 2.74 | 0.38 |
| 3-6 | 0.70 | 0.15 | -0.01 |
| 4-6 | 0.56 | 0.35 | 0.20 |
| 5-6 | 0.07 | 3.52 | 0.34 |
| 1-7 | 0.10 | 2.78 | -0.49 |
| 2-7 | 0.77 | 0.09 | -0.01 |
| 3-7 | 0.07 | 3.40 | 0.46 |
| 4-7 | 0.60 | 0.27 | 0.06 |
| 5-7 | 0.10 | 2.80 | 0.46 |
| 6-7 | 0.06 | 3.79 | 0.40 |
| 1-8 | 0.64 | 0.23 | 0.08 |
| 2-8 | 0.53 | 0.39 | -0.01 |
| 3-8 | 0.13 | 2.39 | 0.25 |
| 4-8 | 0.66 | 0.20 | 0.13 |
| 5-8 | 0.98 | 0.00 | -0.03 |
| 6-8 | 0.56 | 0.34 | 0.10 |
| 7-8 | 0.76 | 0.09 | 0.05 |
| 1-9 | 0.96 | 0.00 | 0.00 |
| 2-9 | 0.12 | 2.50 | 0.25 |
| 3-9 | 0.54 | 0.38 | -0.03 |
| 4-9 | 0.19 | 1.77 | -0.41 |
| 5-9 | 0.43 | 0.63 | 0.29 |
| 6-9 | 0.11 | 2.62 | 0.39 |
| 7-9 | 0.02 | 5.37 | 0.36 |
| 8-9 | 0.71 | 0.14 | 0.08 |
| 1-10 | 0.48 | 0.50 | -0.29 |
| 2-10 | 0.89 | 0.02 | -0.15 |
| 3-10 | 0.99 | 0.00 | 0.01 |
| 4-10 | 0.78 | 0.08 | -0.18 |
| 5-10 | 0.97 | 0.00 | -0.06 |
| 6-10 | 0.22 | 1.58 | -0.37 |
| 7-10 | 0.72 | 0.13 | -0.02 |
| 8-10 | 0.20 | 1.68 | -0.27 |
| 9-10 | 0.56 | 0.34 | 0.10 |
| 1-11 | 0.15 | 2.19 | 0.42 |
| 2-11 | 0.97 | 0.00 | -0.11 |
| 3-11 | 0.92 | 0.01 | -0.03 |
| 4-11 | 0.97 | 0.00 | 0.01 |
| 5-11 | 0.75 | 0.11 | 0.08 |
| 6-11 | 0.32 | 0.99 | -0.37 |
| 7-11 | 0.76 | 0.09 | -0.07 |
| 8-11 | 1.00 | 0.00 | -0.04 |
| 9-11 | 0.66 | 0.19 | 0.06 |
| 10-11 | 0.88 | 0.02 | -0.03 |

Table SI 7: Results of the ANCOVA analysis to assess differences in iCAPs Anti-couplings between HC and patients with PMS. Effect size is reported as Cohen's d.

| iCAPs pair | p value | F(1,52) | effect size (Cohen's d) |
|---|---|---|---|
| Anti-couplings | | | |
| 1-2 | 0.20 | 1.69 | 0.34 |
| 1-3 | 0.43 | 0.64 | 0.18 |
| 2-3 | 0.08 | 3.20 | -0.59 |
| 1-4 | 0.22 | 1.56 | -0.24 |
| 2-4 | 0.48 | 0.50 | -0.12 |
| 3-4 | 0.32 | 1.01 | 0.31 |
| 1-5 | 0.96 | 0.00 | 0.02 |
| 2-5 | 0.34 | 0.93 | 0.34 |
| 3-5 | 0.87 | 0.03 | 0.00 |
| 4-5 | 0.01 | 7.20 | 0.90 |
| 1-6 | 0.47 | 0.53 | 0.02 |
| 2-6 | 0.66 | 0.19 | -0.18 |
| 3-6 | 0.07 | 3.33 | 0.41 |
| 4-6 | 0.82 | 0.05 | -0.17 |
| 5-6 | 0.28 | 1.18 | 0.24 |
| 1-7 | 0.95 | 0.00 | 0.02 |
| 2-7 | 0.25 | 1.37 | 0.22 |
| 3-7 | 0.16 | 2.05 | -0.50 |
| 4-7 | 0.70 | 0.15 | 0.11 |
| 5-7 | 0.79 | 0.07 | 0.13 |
| 6-7 | 0.83 | 0.05 | -0.04 |
| 1-8 | 0.60 | 0.27 | -0.14 |
| 2-8 | 0.23 | 1.48 | -0.35 |
| 3-8 | 0.76 | 0.10 | -0.14 |
| 4-8 | 0.82 | 0.05 | 0.01 |
| 5-8 | 0.71 | 0.14 | 0.07 |
| 6-8 | 0.22 | 1.55 | -0.37 |
| 7-8 | 0.83 | 0.05 | -0.12 |
| 1-9 | 0.64 | 0.22 | 0.05 |
| 2-9 | 0.94 | 0.01 | -0.08 |
| 3-9 | 0.57 | 0.33 | -0.16 |
| 4-9 | 0.08 | 3.27 | 0.48 |
| 5-9 | 0.73 | 0.12 | -0.07 |
| 6-9 | 0.77 | 0.09 | -0.01 |
| 7-9 | 0.36 | 0.86 | -0.19 |
| 8-9 | 0.65 | 0.21 | 0.07 |
| 1-10 | 0.96 | 0.00 | -0.04 |
| 2-10 | 0.44 | 0.61 | -0.22 |
| 3-10 | 0.57 | 0.33 | -0.24 |
| 4-10 | 0.82 | 0.06 | -0.06 |
| 5-10 | 0.38 | 0.80 | -0.31 |
| 6-10 | 0.87 | 0.03 | 0.05 |
| 7-10 | 0.44 | 0.62 | -0.35 |
| 8-10 | 0.50 | 0.46 | -0.34 |
| 9-10 | 0.41 | 0.70 | -0.28 |
| 1-11 | 0.13 | 2.40 | -0.55 |
| 2-11 | 0.41 | 0.70 | 0.20 |
| 3-11 | 0.25 | 1.33 | 0.23 |
| 4-11 | 0.23 | 1.45 | 0.39 |
| 5-11 | 0.74 | 0.12 | -0.06 |
| 6-11 | 0.55 | 0.37 | 0.23 |
| 7-11 | 0.62 | 0.25 | -0.10 |
| 8-11 | 0.11 | 2.65 | 0.38 |
| 9-11 | 0.88 | 0.02 | -0.08 |
| 10-11 | 0.43 | 0.62 | -0.30 |

Table SI 8: Results of the ANCOVA analysis to assess differences in iCAPs duration between CI and CP patients. Effect size is reported as Cohen's d.

| iCAP | p value | F(1,27) | effect size (Cohen's d) |
|---|---|---|---|
| 1. Deep GM | 0.59 | 0.30 | -0.32 |
| 2. Auditory/Sensory-Motor | 0.90 | 0.02 | 0.13 |
| 3. Primary Visual | 0.75 | 0.10 | 0.08 |
| 4. ECN | 0.68 | 0.18 | 0.07 |
| 5. aDMN | 0.23 | 1.54 | -0.83 |
| 6. SAL | 0.80 | 0.07 | -0.08 |
| 7. TempPar/LAN | 0.93 | 0.01 | 0.24 |
| 8. Secondary Visual | 0.11 | 2.76 | -0.32 |
| 9. pCun/pDMN | 0.69 | 0.17 | 0.35 |
| 10. Amy/TempPole | 0.86 | 0.03 | 0.12 |
| 11. OFC | 0.22 | 1.57 | -0.45 |

Table SI 9: Results of the ANCOVA analysis to assess differences in iCAPs Couplings between CI and CP patients with PMS. Effect size is reported as Cohen's d.

| iCAPs pair | p value | F(1,27) | effect size (Cohen's d) |
|---|---|---|---|
| Couplings | | | |
| 1-2 | 0.71 | 0.14 | -0.11 |
| 1-3 | 0.39 | 0.78 | 0.17 |
| 2-3 | 0.42 | 0.67 | 0.47 |
| 1-4 | 0.001 | 13.10 | 1.53 |
| 2-4 | 0.94 | 0.005 | 0.11 |
| 3-4 | 0.13 | 2.46 | -0.39 |
| 1-5 | 0.35 | 0.91 | -0.35 |
| 2-5 | 0.98 | 0.0004 | -0.38 |
| 3-5 | 0.27 | 1.26 | -0.49 |
| 4-5 | 0.76 | 0.09 | -0.10 |
| 1-6 | 0.63 | 0.24 | -0.10 |
| 2-6 | 0.91 | 0.01 | -0.18 |
| 3-6 | 0.89 | 0.02 | -0.13 |
| 4-6 | 0.57 | 0.32 | 0.17 |
| 5-6 | 0.54 | 0.38 | -0.005 |
| 1-7 | 0.47 | 0.53 | -0.29 |
| 2-7 | 0.78 | 0.08 | 0.06 |
| 3-7 | 0.16 | 2.05 | 0.58 |
| 4-7 | 0.26 | 1.3 | -0.34 |
| 5-7 | 0.54 | 0.39 | -0.09 |
| 6-7 | 0.18 | 1.87 | -0.06 |
| 1-8 | 0.83 | 0.04 | -0.20 |
| 2-8 | 0.57 | 0.34 | 0.68 |
| 3-8 | 0.42 | 0.67 | -0.19 |
| 4-8 | 0.72 | 0.14 | 0.11 |
| 5-8 | 0.81 | 0.06 | -0.2 |
| 6-8 | 0.64 | 0.22 | 0.16 |
| 7-8 | 0.27 | 1.25 | 0.02 |
| 1-9 | 0.36 | 0.85 | -0.13 |
| 2-9 | 0.65 | 0.21 | 0.32 |
| 3-9 | 0.70 | 0.15 | -0.33 |
| 4-9 | 0.58 | 0.31 | 0.40 |
| 5-9 | 0.64 | 0.22 | -0.11 |
| 6-9 | 0.15 | 2.19 | 0.64 |
| 7-9 | 0.48 | 0.51 | 0.60 |
| 8-9 | 0.82 | 0.05 | 0.01 |
| 1-10 | 0.65 | 0.21 | -0.10 |
| 2-10 | 0.55 | 0.36 | -0.01 |
| 3-10 | 0.62 | 0.25 | -0.17 |
| 4-10 | 0.55 | 0.37 | -0.22 |
| 5-10 | 0.11 | 2.75 | 0.50 |
| 6-10 | 0.52 | 0.42 | -0.45 |
| 7-10 | 0.22 | 1.6 | 0.24 |
| 8-10 | 0.90 | 0.02 | -0.29 |
| 9-10 | 0.16 | 2.12 | -0.41 |
| 1-11 | 0.55 | 0.37 | 0.08 |
| 2-11 | 0.01 | 8.35 | -0.86 |
| 3-11 | 0.19 | 1.78 | -0.43 |
| 4-11 | 0.53 | 0.41 | 0.24 |
| 5-11 | 0.62 | 0.25 | -0.28 |
| 6-11 | 0.10 | 2.84 | -0.56 |
| 7-11 | 0.47 | 0.54 | -0.33 |
| 8-11 | 0.29 | 1.16 | -0.32 |
| 9-11 | 0.65 | 0.21 | -0.08 |
| 10-11 | 0.76 | 0.10 | 0.05 |

Table SI 10: Results of the ANCOVA analysis to assess differences in iCAPs Anti-Couplings between CI and CP patients with PMS. Effect size is reported as Cohen's d.

| iCAPs pair | p value | F(1,27) | effect size (Cohen's d) |
|---|---|---|---|
| Anti-Couplings | | | |
| 1-2 | 0.80 | 0.06 | 0.04 |
| 1-3 | 0.44 | 0.62 | -0.25 |
| 2-3 | 0.11 | 2.75 | -0.53 |
| 1-4 | 0.03 | 5.22 | -1.11 |
| 2-4 | 0.47 | 0.54 | 0.41 |
| 3-4 | 0.01 | 6.93 | 1.12 |
| 1-5 | 0.65 | 0.21 | -0.17 |
| 2-5 | 0.38 | 0.81 | -0.22 |
| 3-5 | 0.98 | 0.00 | -0.07 |
| 4-5 | 0.58 | 0.31 | 0.04 |
| 1-6 | 0.12 | 2.59 | 0.57 |
| 2-6 | 0.74 | 0.12 | -0.06 |
| 3-6 | 0.87 | 0.03 | 0.02 |
| 4-6 | 0.21 | 1.63 | 0.17 |
| 5-6 | 0.79 | 0.08 | -0.04 |
| 1-7 | 0.39 | 0.77 | 0.39 |
| 2-7 | 0.47 | 0.53 | 0.46 |
| 3-7 | 0.24 | 1.47 | -0.41 |
| 4-7 | 0.12 | 2.56 | 0.78 |
| 5-7 | 0.64 | 0.22 | -0.45 |
| 6-7 | 0.27 | 1.24 | 0.74 |
| 1-8 | 0.51 | 0.45 | -0.16 |
| 2-8 | 0.23 | 1.50 | -0.59 |
| 3-8 | 0.34 | 0.96 | -0.06 |
| 4-8 | 0.62 | 0.26 | -0.09 |
| 5-8 | 0.83 | 0.05 | 0.18 |
| 6-8 | 0.08 | 3.31 | -0.59 |
| 7-8 | 0.48 | 0.52 | 0.14 |
| 1-9 | 0.15 | 2.20 | 0.21 |
| 2-9 | 0.63 | 0.23 | -0.03 |
| 3-9 | 0.74 | 0.11 | 0.28 |
| 4-9 | 0.43 | 0.65 | 0.47 |
| 5-9 | 0.19 | 1.80 | 0.38 |
| 6-9 | 0.27 | 1.29 | -0.54 |
| 7-9 | 0.72 | 0.13 | 0.09 |
| 8-9 | 0.28 | 1.24 | -0.14 |
| 1-10 | 0.64 | 0.23 | -0.18 |
| 2-10 | 0.37 | 0.84 | 0.13 |
| 3-10 | 0.87 | 0.03 | 0.09 |
| 4-10 | 0.29 | 1.15 | 0.42 |
| 5-10 | 0.15 | 2.19 | -0.50 |
| 6-10 | 0.28 | 1.20 | 0.40 |
| 7-10 | 0.39 | 0.78 | 0.02 |
| 8-10 | 0.83 | 0.05 | 0.19 |
| 9-10 | 0.16 | 2.08 | 0.56 |
| 1-11 | 0.76 | 0.09 | -0.05 |
| 2-11 | 0.64 | 0.22 | 0.12 |
| 3-11 | 0.63 | 0.23 | -0.16 |
| 4-11 | 0.92 | 0.01 | 0.13 |
| 5-11 | 0.19 | 1.78 | -0.48 |
| 6-11 | 0.81 | 0.06 | -0.17 |
| 7-11 | 0.99 | 0.00 | 0.07 |
| 8-11 | 0.30 | 1.10 | -0.16 |
| 9-11 | 0.40 | 0.73 | -0.09 |
| 10-11 | 0.51 | 0.44 | 0.11 |

Figure SI 4: Bar plot representing the 11 iCAPs duration (A) and the percentage of opposite (anti-couplings. B) or same (couplings. C) sign overlap between iCAP 5 and the other iCAPs for HC and MS patients at baseline (subsample of 35 subjects) and FU. $*p < 0.05$, $**p < 0.01$.

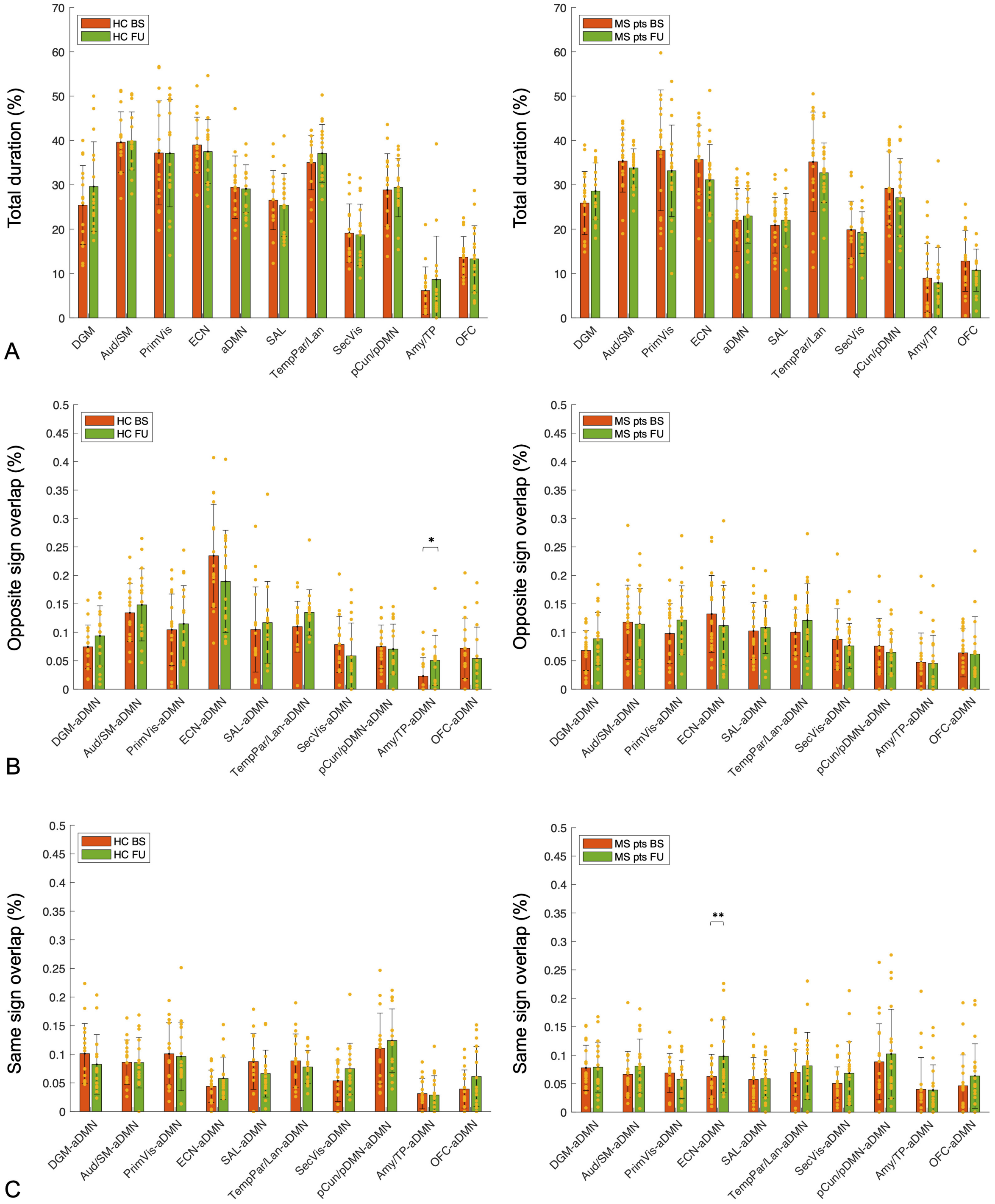

Figure SI 5: Grouped PLSC between aDMN anti-couplings and cognitive test scores. The anti-couplings explain the behavioral variance in the CI group. Specifically, the SDMT and BVMT-R were inversely associated to the aDMN-Aud/SM and aDMN-pCun/pDMN anti-coupling; the SDMT additionally correlated positively with aDMN-SecVis and negatively with aDMN-ECN anti-couplings. Instead, CVLT-II was inversely associated to the aDMN-Amy/TP anti-coupling but positively to the aDMN-Aud/SM and aDMN/SAL anti-couplings

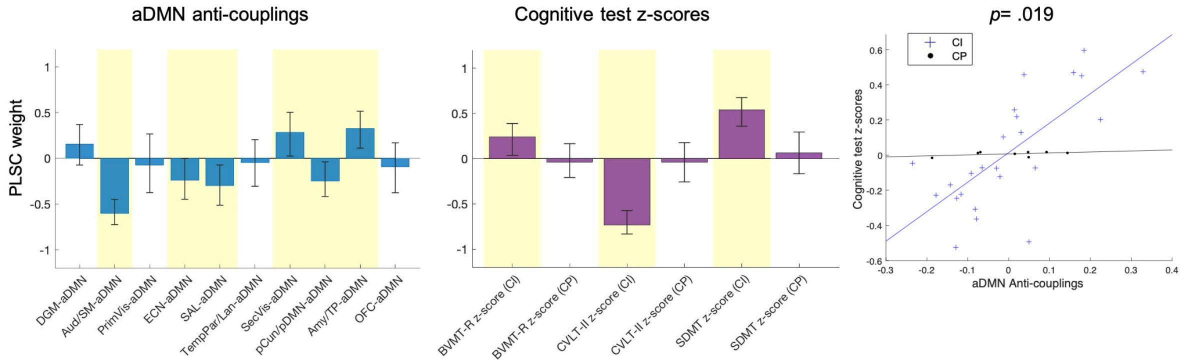


Figure SI 6: PLSC between all iCAPs durations and clinical test scores. In particualr, the EDSS and T25FW test positively correlated to the DGM and inversely to Aud/SM and pCun/pDMN durations. Moreover, T25FW inversely correlated with the TempPar/LAN duration. It emerges again the association between aDMN and cognitive test scores.

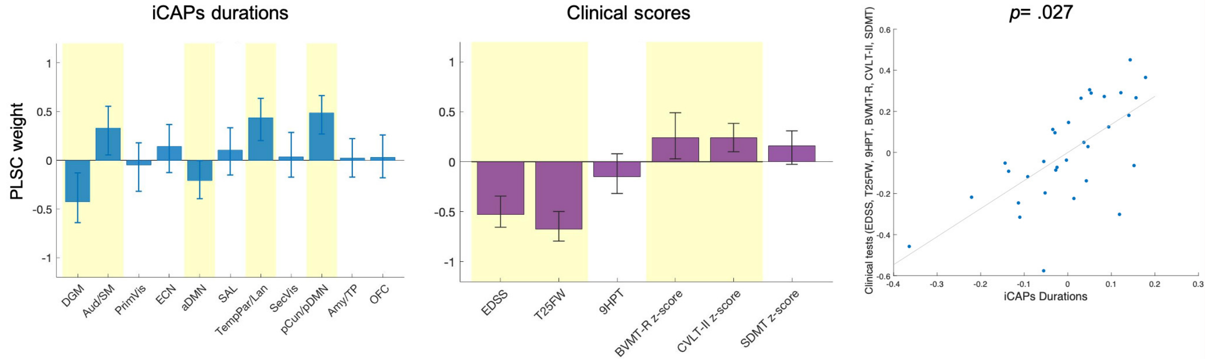

Figure SI 7: Spatial patterns of the 11 iCAPs retrieved from the analysis on the subsample of 35 HC and patients with PMS. Under each iCAP the average consensus and the number of innovation frames assigned to it. Coordinates refer to the Montreal Neurological Institute space. Each iCAP is numbered as the correspondent resulted from the original analysis on 57 subjects. DGM: deep grey matter; Aud/SM: the auditory/sensory-motor; PrimVis: primary visual; ECN: executive control network; aDMN: anterior DMN; SAL: salience; TempPar/Lan: temporo-parietal/language; SecVis: secondary visual; pCun/pDMN: precuneus/posterior DMN; Amy/TP: amygdala/temporal pole; OFC:orbito-frontal cortex.

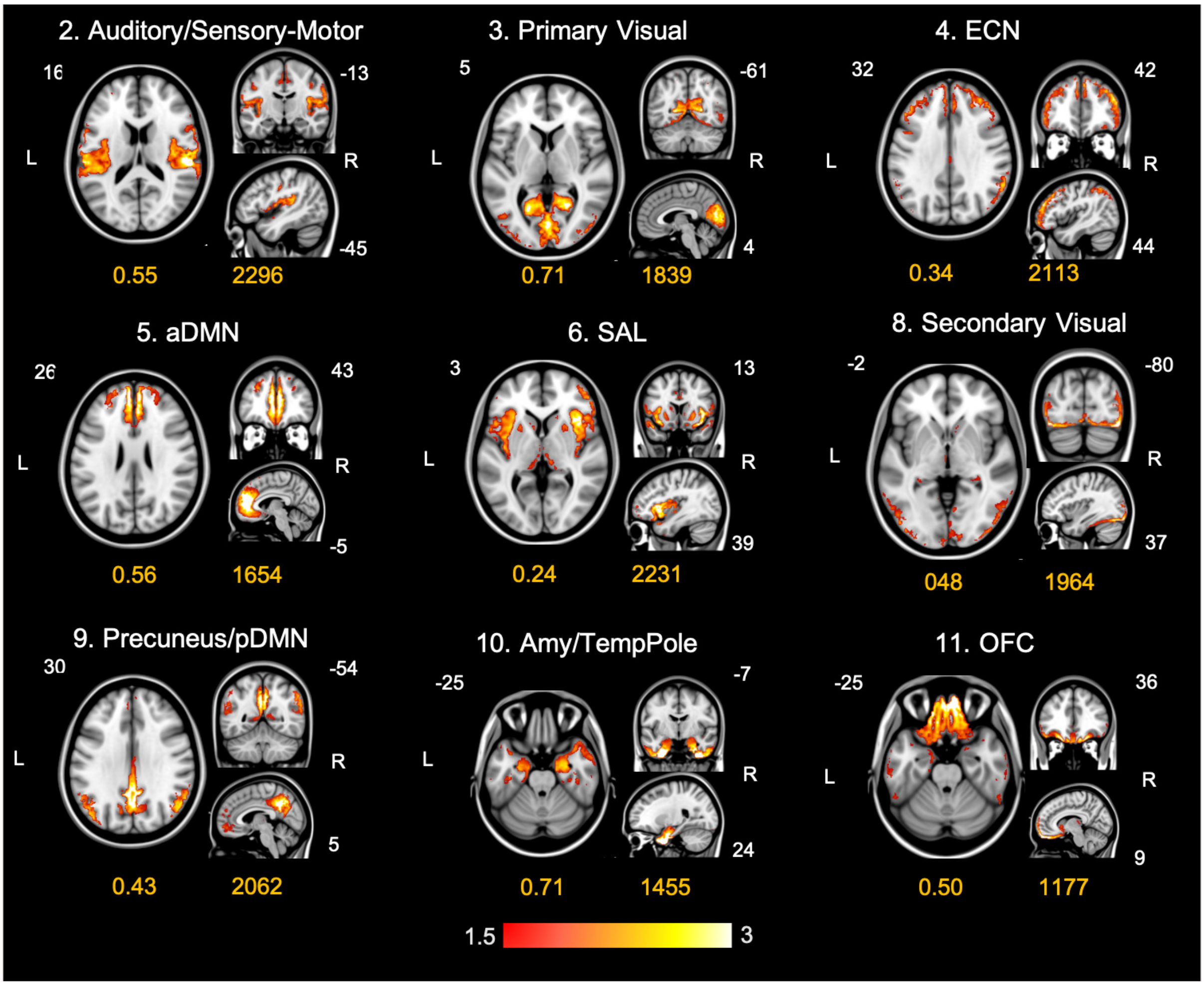

Table SI 11: Appendix for MRI methodology at baseline and FU.

| | |
|---|---|
| Field strength | 3.0 T |
| Manufacturer | Siemens |
| Model | MAGNETOM Skyra syngo MR D13 |
| Coil type | head coil |
| Number of coil channels | 32 channel |
| Type | resting-state fMRI, 3D T1 MPRAGE, T2–weighted TSE sequence |
| Acquisition time | resting-state fMRI: 6.51 min |
| | 3D T1 MPRAGE: 8.45 min |
| | T2–weighted TSE: 2.42 min |
| Orientation | resting-state fMRI: T ¿ C-20.0 |
| | 3D T1 MPRAGE: Sagittal |
| | T2–weighted TSE: Transversal |
| Voxel size | resting-state fMRI: 2.1×2.1×2.1 mm |
| | 3D T1 MPRAGE: 0.8×0.8×0.8 mm |
| | T2–weighted TSE: 0.5×0.5×3.0 mm |
| TR | resting-state fMRI: 1000 ms |
| | 3D T1 MPRAGE: 3000.0 ms |
| | T2–weighted TSE: 8000.0 ms |
| TE | resting-state fMRI: 35.00 ms |
| | 3D T1 MPRAGE: 2.47 ms |
| | T2–weighted TSE: 95.00 ms |
| TI | 3D T1 MPRAGE: 1000 ms |
| Flip angle | resting-state fMRI: 60 deg |
| | 3D T1 MPRAGE: 7 deg |
| | T2–weighted TSE: 160 deg |
| Field of view | resting-state fMRI: 228x228 mm2 |
| | 3D T1 MPRAGE: 256x256 mm2 |
| | T2–weighted TSE: 256x256 mm2 |
| Parallel imaging | resting-state fMRI: none |
| | 3D T1 MPRAGE: GRAPPA |
| | T2–weighted TSE:none |
| Contrast enhancement | No |
| Lesions | |
| Type | T2-hyperintense, T1-hypointense |
| Analysis method | Semiautomatic segmentation technique |
| Analysis software | Jim software package (Version 7, , Xinapse Systems, Northants, UK) |
| Output measure | Lesion volume [ml] |
| Tissue volumes | |
| Type | Whole brain, gray matter and white matter normalized volumes, percentage of brain volume changes at follow-up |
| Analysis method | Non-brain tissue stripping, brain segmentation and skull based normalization |
| Analysis software | SIENAX SIENA |
| Output measure | Normalized brain volume (NBV), normalized grey matter volume, normalized white matter volume. Percentage brain of volume change (PBVC) |
| Other MRI measures | |
| Type | RS fMRI dynamic functional activity |
| Analysis method | RS fMRI pre-processing iCAPs pipeline (https://c4science.ch/source/iCAPs/) Calculation of the temporal properties of iCAPs (overall duration and couplings/anti-couplings between each pair of iCAPs |
| Analysis software | SPM12, MATLAB, DPARSF, IBASPM |
| Output measure | Overall duration of each iCAP (or brain state, retrieved from innovation signals, which encode onsets of activations/de-activations by positive/ negative spikes,) and interactions between each pair of iCAPs. |
